# Melanoma to rhabdomyosarcoma plasticity in the setting of immunotherapy

**DOI:** 10.1101/2025.10.31.25338685

**Authors:** Andrew D. Knight, Emily J. Robitschek, Jia-Ren Lin, Dennie T. Frederick, Alvin Shi, Ana B. Larque, Benchun Miao, Rumya S. Raghavan, Tatyana Sharova, John H. Shin, Peter Sorger, Manolis Kellis, Keith T. Flaherty, Nir Hacohen, Genevieve M. Boland, Ivan Chebib, David Liu, Ryan J. Sullivan, Arnav Mehta

## Abstract

Acquired resistance to immune checkpoint inhibitors (ICIs) remains a significant challenge in the treatment of metastatic melanoma. Phenotypic plasticity, such as dedifferentiation and transdifferentiation, is an increasingly recognized mechanism for treatment resistance. We present a case of a man in his 70s with metastatic melanoma who experienced progression through sequential treatments including pembrolizumab in combination with the HDAC inhibitor entinostat, and ipilimumab. During treatment a histologically distinct pleomorphic rhabdomyosarcoma (RMS) emerged at metastatic sites. Longitudinally acquired tumor samples representing both phenotypes were analyzed using whole-exome sequencing (WES), RNA sequencing (RNA-seq) and high-plex tissue imaging (spatial proteomics). WES revealed driver mutations (e.g. NRAS, NF1) and loss-of-heterozygosity (LOH) shared between phenotypes indicating a common ancestral clone. Phylogenetic analysis demonstrated an early divergence of the phenotypes, with each later acquiring unique mutations. RNA-seq showed mutually exclusive expression of lineage-specific markers as well as epithelial-mesenchymal transition and myogenic gene set enrichment in the RMS samples. High-plex imaging identified distinct tumor microenvironments, with RMS lesions enriched in CD163^+^ macrophages.

**Statement of Significance:** Transdifferentiation has been observed in a wide range of malignancies, but the molecular mechanisms of this phenomenon are poorly understood. This case provides the first molecularly validated example of melanoma to rhabdomyosarcoma transdifferentiation presenting as spatially segregated metastatic lesions with distinct, unmixed histologies and illustrates a mechanism of resistance to immunotherapy driven by phenotypic plasticity.

## Introduction

Immune checkpoint inhibitors (ICIs) have significantly improved outcomes for patients with metastatic melanoma with many experiencing durable benefit.^1^ However, a substantial subset of patients have primary resistance or develop secondary resistance. Understanding the mechanisms of resistance is important for developing strategies to expand the benefit of these transformative therapies. Resistance mechanisms to ICIs are diverse and include tumor extrinsic factors such as an immunosuppressive tumor microenvironment as well as tumor intrinsic factors such as defects in antigen presentation and low antigenicity, including by MHC class I loss of heterozygosity, tumor intrinsic mutations that confer resistance to T cell killing.^2^ In addition, phenotypic plasticity of tumor cells has been increasingly recognized as a driver of therapeutic resistance.^3^ Phenotypic plasticity can manifest as dedifferentiation, as has been seen for melanoma in the setting of targeted therapy and immunotherapy, or more dramatically through lineage switching or transdifferentiation, where tumor cells adopt features seen in tissues of distinct embryologic origin, as has been observed in adenocarcinoma to neuroendocrine transformation in prostate and lung cancers.^4–6^

Melanoma is known for its phenotypic heterogeneity, which frequently includes dedifferentiation, characterized by the loss of canonical markers. In contrast, a complete lineage switch to a different, differentiated cell type known as transdifferentiation has been only rarely reported. Prior reports described co-existing melanoma and sarcoma-like phenotypes lacked definitive molecular evidence to indicate a shared clonal origin or identify the evolutionary trajectory while on treatment.^7–9^ Here we present a unique case of metastatic melanoma that developed acquired resistance to ICI treatment. During treatment, biopsies identified lesions exhibiting a pleomorphic rhabdomyosarcoma (RMS) phenotype alongside persistent melanoma. Utilizing longitudinal multiomic profiling we demonstrate the shared clonal origin of these histologically distinct tumors providing evidence for phenotypic plasticity as a mechanism of immunotherapy resistance in patients.

## Results

### Case Presentation

A man in his 70s presented with metastatic melanoma following a history of previously excised primary melanomas. At the time of the current presentation, he developed a flesh-colored lesion that grew over a 3-week period on his left shoulder. Biopsy revealed a superficial spreading, ulcerated melanoma (1.75mm depth, 6 mitoses/mm²). Wide excision and SLNB were performed with metastatic involvement identified in a single left axillary lymph node (Sample MEL_LN; **Figure 1A**). A completion lymphadenectomy revealed no further nodal involvement. Staging PET/CT showed abnormal uptake only in the thyroid. A thyroidectomy shortly thereafter showed no evidence of cancer. He was therefore diagnosed with a stage IIIA melanoma.

**Figure 1:**
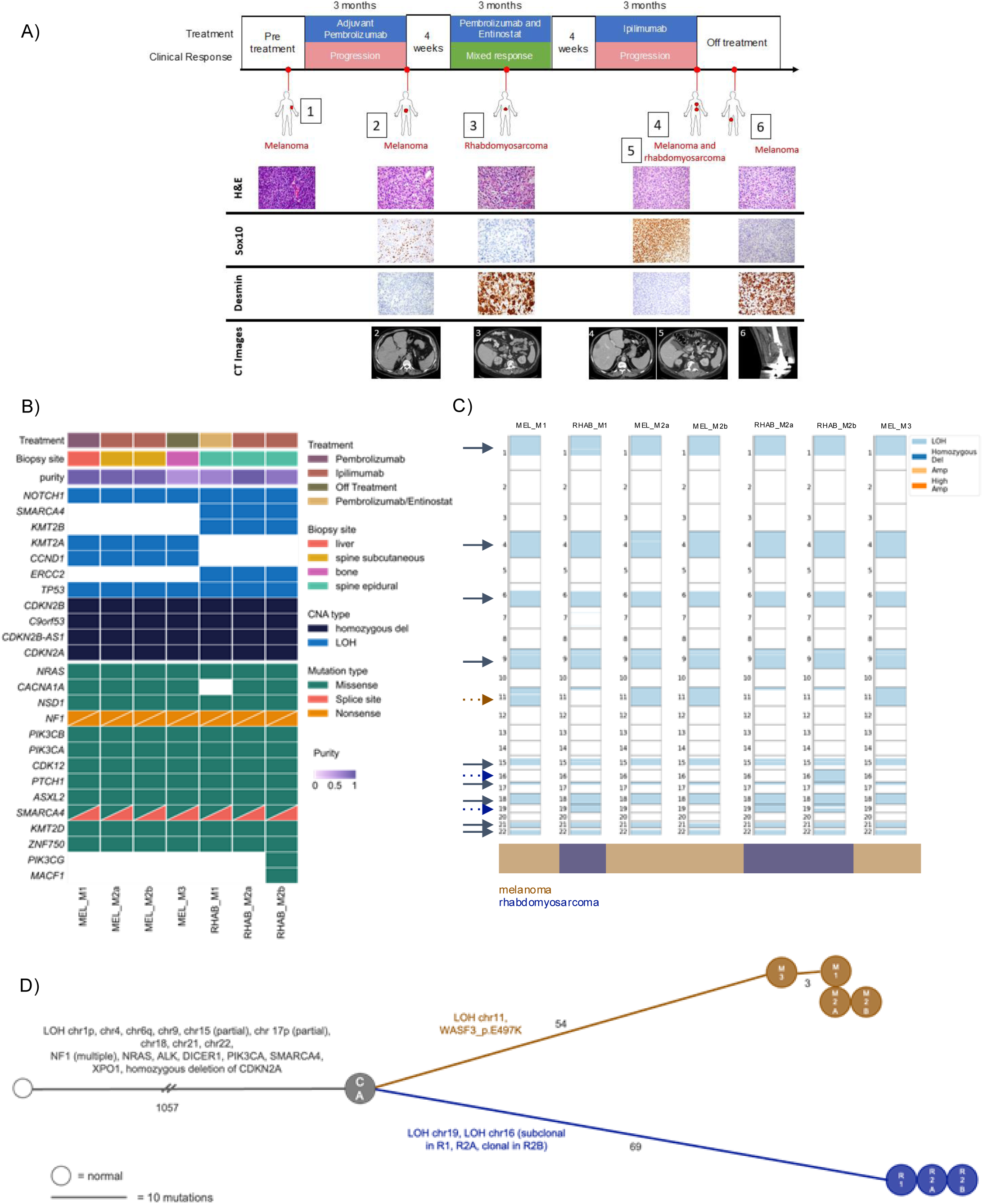
The rhabdomyosarcoma and melanoma tumors share many genomic features and arose from the same common ancestor within the patient. A) Clinical course for the patient, including relative times at which the tumors were detected and sampled for WES and H&E showing the cellular morphology (H&E) and staining for a common melanoma marker (Sox10), and for a common rhabdomyosarcoma marker (Desmin). B) Comut plot showing clinical metadata (treatment, biopsy site) and tumor purity for the samples alongside some of the driver mutations and copy number alterations in these tumors and how these are largely shared across samples. C) Plot of the copy number alterations across the samples, including those shared across all samples (gray arrows), uniquely shared across melanoma samples (dashed brown arrow), and those uniquely shared across rhabdomyosarcoma samples (dashed blue arrow). The LOH in chr16 is subclonal in RHAB_M1 and RHAB_M2a so it is only colored here in RHAB_M2b where it is clonal. D) Plot of lineage tree summarizing large scale CNA changes and mutational differences (from pyclone) shared by all tumors, melanoma tumors (brown), and rhabdomyosarcoma tumors (blue) with key driver mutations labeled. “M” refers to the melanoma lineage and “R” refers to the rhabdomyosarcoma phenotype lineage.

The patient was enrolled in a clinical trial, which randomized patients to pembrolizumab versus the investigator’s choice of ipilimumab or high-dose interferon. He was randomized to receive adjuvant pembrolizumab group (200mg every 3 weeks). After four cycles, a re-staging PET/CT scan showed multiple new liver and bone lesions. A liver biopsy was obtained (Sample MEL_M1; **Figure 1A**). Immunohistochemistry (IHC) staining was positive for S100, MART-1, SOX10 and MITF consistent with metastatic melanoma. BRAF V600E IHC was negative. Importantly, 80% of the tumor cells had strong staining for PD-L1 expression and CD8^+^ T-cell infiltration was associated with 25% of the tumor.

Given the progression on pembrolizumab, he was started on a phase 1b/2 trial of entinostat, a histone deacetylase (HDAC) inhibitor, in combination with pembrolizumab. Per study protocol, he received 5mg of entinostat by mouth weekly and 200mg of pembrolizumab by intravenous infusion every three weeks. A re-staging PET/CT one month after initiation of the trial revealed a mixed response with interval regression of his liver metastases, but several new lytic lesions in the spine. Shortly thereafter, he developed severe back pain and underwent a resection of an L2 epidural and vertebral body tumor along with a decompression of his L2 and L3 nerve roots (Sample RHAB_M1; **Figure 1A**). Pathology of this tumor identified a distinct histologic entity from his melanoma with features characteristic of pleomorphic rhabdomyosarcoma. The tumor stained diffusely positive for desmin and MyoD1, and had multifocal positivity for myogenin. The tumor stained negative for S100, SOX10, MNF116, EMA and MDM2. There was no PD-L1 staining and less than 5% tumor CD8^+^ T-cell infiltration.

Three weeks after surgery the patient re-presented with severe back pain and underwent stereotactic radiosurgery to the L2 tumor bed. A follow-up PET/CT scan revealed progression of several tumors, including his previously regressed liver metastases. Entinostat and pembrolizumab were discontinued due to progression of disease. He was started on ipilimumab 3mg/kg every three weeks. After four cycles, follow-up imaging revealed multifocal progression of disease. He again developed worsening back pain due to progression spinal metastases and underwent an L1-L3 revision, decompression, and resection of an L1-L2 epidural tumor and T12-L1 subcutaneous tumor. Pathology confirmed the presence of two distinct phenotypes. The L1-L2 epidural tumor remained consistent with rhabdomyosarcoma, staining positive for desmin, myogenin and myoD1, and negative for the melanoma markers S100, MITF and HMB-45 (Sample RHAB_M2; **Figure 1A**). The T12-L1 subcutaneous tumor was consistent with metastatic melanoma, exhibiting spindle cell morphology with positive S100 and MITF staining, patchy MITF and rare HMB-45 staining, and no desmin, myogenin and myoD1 staining (Sample MEL_M2; **Figure 1A**).

While recovering from surgery, the patient developed a femoral metastatic lesion. A sample was obtained from this lesion and identified as a malignant spindle cell neoplasm consistent with metastatic melanoma (MEL_M3; **Figure1A**). The tumor stained positive for S100, SOX10, MART-1 (rare cells) and negative for desmin and myogenin. The patient was unable to pursue additional systemic therapy due to poor performance status at this point and ultimately succumbed to his metastatic disease. The patient’s treatment course and biopsies are summarized in **Figure 1A**.

### Genomic sequencing reveals a common origin for melanoma and rhabdomyosarcoma lesions

Whole exome sequencing was performed on seven longitudinally collected metastatic specimens to investigate the relationship between RMS and melanoma lesions (**Figure 1A and Supplementary Table 1**). Sequenced tumors shared 954 SNV (single nucleotide variant) mutations, which represented the vast majority (82%) of the total 1,170 mutations called across all the tumor samples. We additionally saw common clonal loss of heterozygosity (LOH) of 1p, 4, 6q, 9, 18, 21q, and 22q, and inferred a common ancestor for all the tumors with homozygous deletion of CDKN2A, and driver mutations in NRAS, NF1 (bi-allelic), CDK12, and SMARCA4. Genomic features unique to the melanoma lesions included a clonal LOH of chromosome 11, while RMS lesions shared clonal LOH of chromosome 19 (**Figure 1B-D**). Phylogenetic analysis revealed an early split between histologic melanoma and RMS subtypes (**Figure 1D**). Many of these genomic features (mutations leading to activation of RAS pathway, loss of CDKN2A) were previously reported in the literature characterizing molecular phenotypes in both melanoma and RMS, though largely represent canonical melanoma drivers.^10–12^ Together these data strongly supported the assertion that these tumors derived from a common ancestor, likely of melanoma origin.

There were no mutational or copy number gains exclusive to the RMS lineage that would have pointed to specific genomic features for driver genes causing the phenotype switch. We were thus left with one of three possible hypotheses: (1) loss of genes on chromosome 11 prevented phenotypic transformation from melanoma to RMS; (2) loss of genes in chr19 or chr16 enabled the phenotypic transformation; or (3) the phenotypic transformation was independent of genetic events, and was enabled by epigenetic and transcriptional changes within the melanoma cell. To better dissect these putative drivers of phenotypic transformation, we next sought to study the transcriptional profile of each of these tumors by bulk RNA-sequencing (RNA-seq). Due to sample limitations, we were unable to perform epigenetic profiling.

### Melanoma and rhabdomyosarcoma tumors express distinct, lineage-specific gene programs

RNA-seq analysis of melanoma and rhabdomyosarcoma samples demonstrated dramatically different gene expression profiles between the distinct histologies despite the genetic similarities. Differential gene expression analysis revealed patterns consistent with the distinct lineages observed histologically. Expression of SOX10 and PMEL, commonly used markers in melanoma, were consistently higher on average across the melanoma phenotype samples than the RMS phenotype samples (**Figure 2A and Supplementary Table 2**). In the RMS samples, canonical skeletal muscle differentiation gene expression was increased including DES, MYLA4, MYOG, MYOD1, MYL1, MYL4, TNNT1 and TNNT2 consistent with what we would expect in the RMS phenotype. Gene expression related to melanosome function, such as GPR143 and SLC23A5, was notably higher in the subcutaneous metastatic lesions compared to the RMS samples.^13,14^ XAGE1 genes (XAGE1B and XAGE1C), whose over-expression has previously been reported in rhabdomyosarcomas, were also more highly expressed in rhabdomyosarcoma lesions.^15^ Expression of immune-related genes like IL32 and CHI3L1 were noted to be higher on average for RMS samples, and importantly, so were the expression of IFN-related genes such as ISG15, IFI6 and IFI44L (**Figure 2A**). It remains to be determined what the underlying source of these immune genes may be, which could not be de-convolved using the bulk RNA-seq data. These data may suggest, however, that differences in immune microenvironments could also have played a role in the phenotype transition or may be a consequence of phenotypic transformation, and subsequently play a role in resistance.

**Figure 2:**
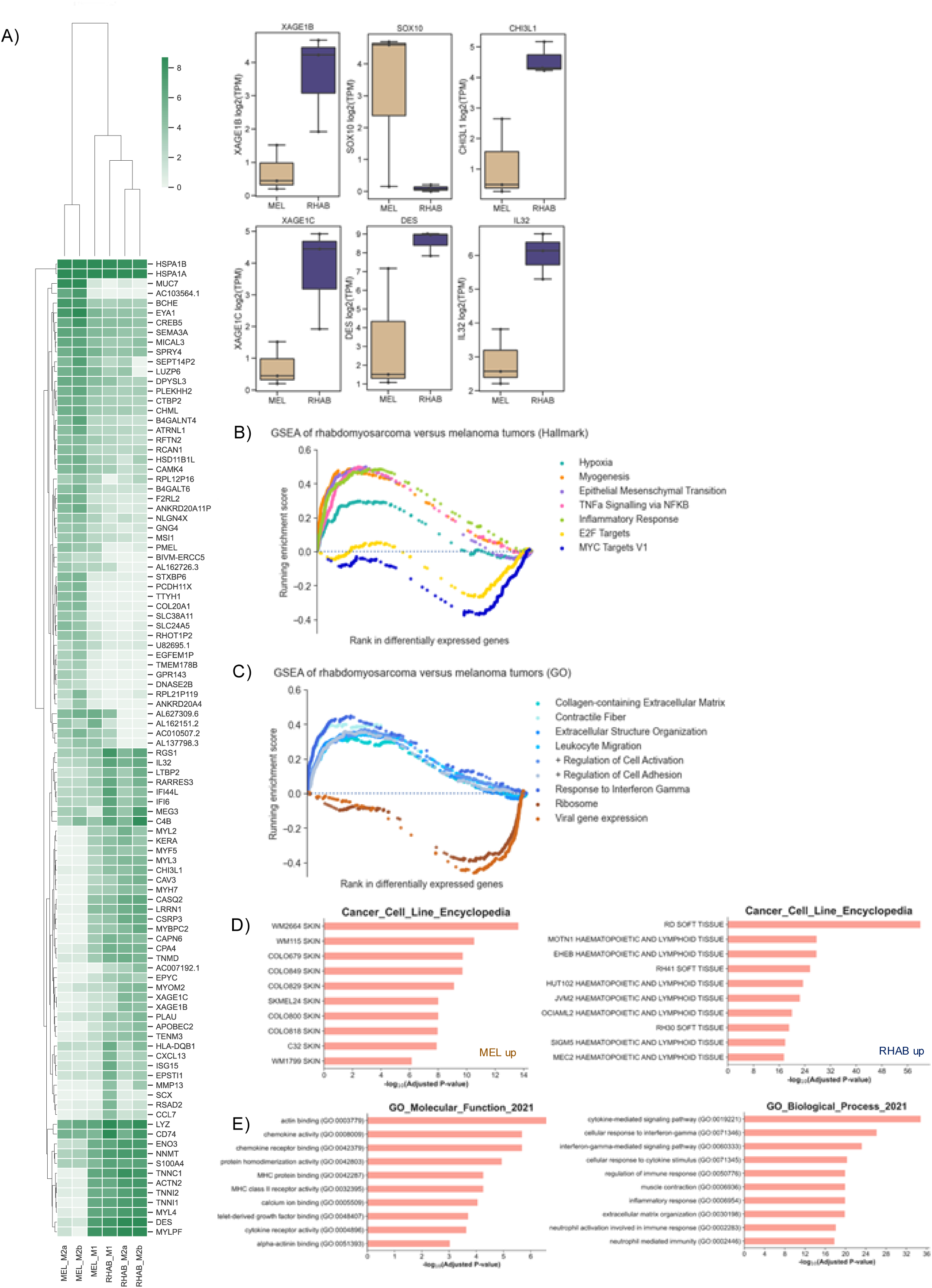
The rhabdomyosarcoma and melanoma tumors are characterized by different patterns of gene expression. A) Top 50 genes upregulated in melanoma samples versus rhabdomyosarcoma samples and vice versa. Selected genes were chosen after the data was ranked by log2FC and filtered by p<0.05 based on a two-sided t test (without multiple hypothesis correction) B-C) Results showing selected results from GSEA of genes ranked by log2FC between rhabdomyosarcoma phenotype tumors and melanoma phenotype tumors (all pathways shown significant with an FDR<0.001). D) Most significantly enriched Cancer Cell Line Encyclopedia (CCLE) GSEA results (top, enriched in melanoma on left and rhabdomyosarcoma phenotype on right). E) GO Molecular Function and GO Biological process pathways in rhabdomyosarcoma phenotype tumors as compared to melanoma phenotype tumors (results from Enricher with top 3000 genes ranked by log2FC for each phenotype)

Interestingly, the transcriptomic profile of the initially biopsied liver metastasis (MEL_M1) revealed an intermediate state between the melanoma and RMS phenotypes. Although the lesion retained expression of melanocytic markers like PMEL, it shared elevated expression of muscle differentiation genes such as DES, MYL4 and MYLPF and decreased expression of genes related to melanosome function, consistent with the RMS samples. These transcriptional changes suggest that rhabdomyosarcoma gene program may have been expressed prior to histological transformation, and we posit that, at the gene expression level, transdifferentiation of lesions from melanoma to rhabdomyosarcoma may occur across a transcriptional spectrum that underlies a continuum of cell states.

Gene Set Enrichment Analysis (GSEA) using genes ranked by differential expression between RMS and melanoma phenotypes identified significant differences in 19 hallmark gene sets (p-value <0.03 and FDR < 0.01). RMS samples were highly enriched for signatures related to myogenesis and epithelial mesenchymal transition (EMT) (**Figure 2B**), which suggests the potential for these tumors to co-opt developmental programs. Several immune signatures were enriched in RMS samples relative to melanoma samples, including inflammatory response and TNF-alpha signaling via NFkB (**Figure 2B,C**). We subsequently used EnrichR to compare transcriptional enrichment of known lineage-specific gene expression programs within each histologic phenotype. Results confirmed enrichment of muscle function GO terms and sarcoma cell line signatures in the RMS phenotype, contrasting with skin cancer cell line signatures enriched in the melanoma phenotype and further reinforced the distinct biological states of these two lineages (**Figure 2D,E**).

### Highly multiplexed tissue imaging reveals distinct immune microenvironments and balance of EGFR and NGFR expressing tumor cells across histologies

To investigate the cellular composition and spatial relationships of the tumors and their microenvironments, we performed highly multiplexed tissue imaging cyclic immunofluorescence; (t-CyCIF) of all tumors for which we had remaining tissue available.^16^ We initially used samples MEL_2a and RHAB_M2a as two representative lesions for melanoma and rhabdomyosarcoma histologies collected at same time when the patient’s spinal tumors were resected. There can be some overlap between melanoma and RMS IHC markers, including vimentin and S100, which could potentially confound analysis in this case.^17,18^ Thus we first stained with markers of melanoma (SOX10, S100, MART1) and sarcoma/mesenchymal (Desmin, Vimentin, Myogenin) to determine if there was either a mixed composition of melanoma and rhabdomyosarcoma tumor cells, or if there was mixed expression in tumor cells at the single cell level. We observed small numbers of S100^+^ and Vimentin^+^ cells in both specimens (**Figure 3A**). However, melanoma (SOX10^+^) cells and RMS (Desmin^+^) cells occupied mutually exclusive territories, demonstrating phenotypic stability and lack of significant intermixing (**Figure 3A-C**). We clustered cells based on marker expression, generating six clusters: RMS (Rhab), melanoma (Mel), stroma, lymphoid (Lymph), macrophage (Macro) and other. Analyzing the samples obtained over the course of treatment supported the conclusion that individual lesions contained cells expressing melanoma markers and cells expressing RMS markers, but no lesions had mixed phenotypes detected (**Figure 3B,C**).

**Figure 3:**
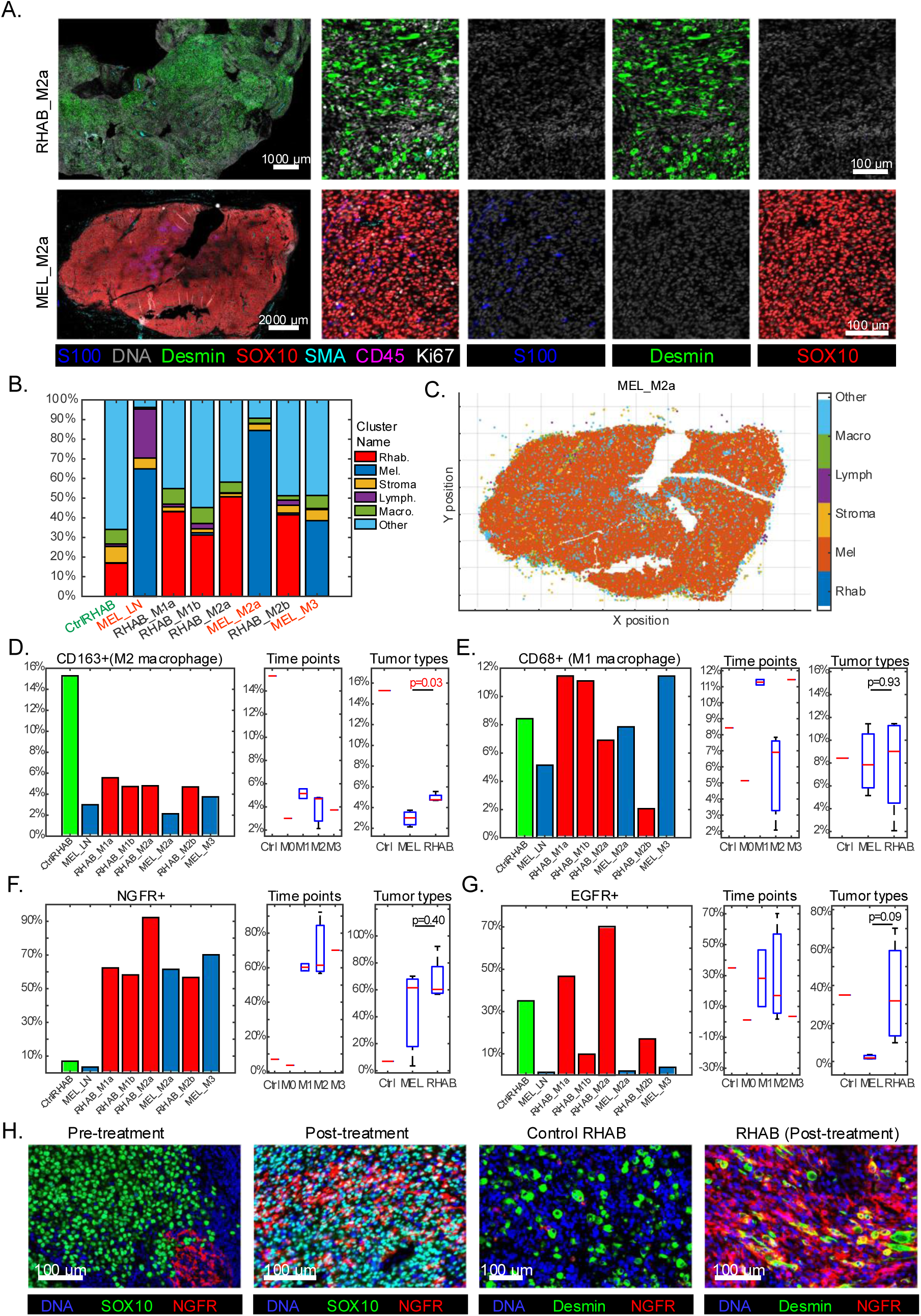
Rhabdomyosarcoma and melanoma tumors are not mixed in phenotype, and are characterized by different patterns of NGFR and EGFR expression and distinct immune microenvironments. A. Example images from Rhabdomyosarcoma (T4_A1) and Melanoma (T4_B3). Tumor (SOX10, S100 and Desmin), immune (CD45) and stromal (SMA) markers are selected B. Distribution of different cell types in each samples. Single-cell clustering was performed with Gaussian Mixture Model (GMM), and 6 main cell types (Rhab, Mel, Stroma, Lymph, Macro, Other) were identified. CasePR2 is the control Rhabdomyosarcoma sample from another patient. The melanoma samples are colored with red and Rhabdomyosarcoma are labeled black. C. The example spatial map of cell types. D. M2 Macrophage (CD163+) fractions in different samples. Left panel: individual sample counts, middle panel: counts by timepoint, right panel: counts by tumor types. E. M1 Macrophage (CD68+) fractions in different samples. Left panel: individual sample counts, middle panel: counts by timepoint, right panel: counts by tumor types. F. NGFR+ cell fractions in different samples. Left panel: individual sample counts, middle panel: counts by timepoint, right panel: counts by tumor types. G. EGFR+ cell fractions in different samples. Left panel: individual sample counts, middle panel: counts by timepoint, right panel: counts by tumor types. H. Induction of NGFR after treatment

We also quantified the immune composition and degree of tumor infiltration in these specimens. While lymphocyte infiltration was generally low, aside from the lymph node specimen (MEL_LN), macrophages were present in both tumor types (Figure 3B). Notably, RMS lesions (including a true RMS from a different patient as a control) contained a significantly higher proportion of the types of macrophages (CD68+/CD163+) that are generally thought to be immunosuppressive as compared to melanoma lesions (**Figure 3D**), while total CD68+ macrophage levels were comparable (**Figure 3E**).

Finally, we sought to evaluate the levels of other signaling pathways or de-differentiation programs. Expression of nerve growth factor receptor (NGFR), a neural crest stem cell marker, is associated with a cancer stem cell-like state in melanoma known to promote metastases and a more aggressive phenotype.^19^ We observed a marked increase in the fraction of NGFR-positive cells in the post-treatment samples, both melanoma and RMS, from 10% to over 50% (**Figure 3F,H**). EGFR expression, commonly seen in RMS, was present in a higher fraction of cells in all histologically RMS samples, though this did not reach statistical significance given the small number of specimens this limited cohort (p = 0.09, **Figure 3G**).^20^

## Discussion

This report describes a striking example of phenotypic plasticity originating in a patient with metastatic melanoma treated with immune checkpoint inhibition. Using genomic sequencing we demonstrate that the two morphologically distinct phenotypes identified in this case shared an ancestral clonal. Whole exome sequencing revealed that 82% of all identified mutations were shared across all samples, including both RMS and melanoma histologies. Importantly, across both tumor types, several canonical melanoma driver alterations (NRAS, NF1, and CDKN2A deletion) were identified, suggesting a shared lineage. The genomic alterations unique to the RMS phenotype were notably lacking activating mutations that would suggest a clear oncogenic driver to explain the lineage switch. The subclonal nature of these genetic alterations, present in only some of the RMS samples, further indicates that these genetic alterations are not necessary for the transition to the RMS phenotype. Instead, the lineage-specific LOH events in the RMS phenotype appear to mark evolutionary divergence whose functional role remains unclear.

Beyond canonical melanoma mutations, the presence of a shared, clonal mutation in SMARCA4 is particularly noteworthy. SMARCA4, also known as BRG1, is a component of the SWI/SNF chromatin remodeling complex, which alters nucleosome positioning to regulate gene expression.^21^ Under normal physiologic conditions, SMARCA4 is important for maintaining lineage identity in multiple tumor types.^22^ Loss of SMARCA4 activity has been implicated in tumor dedifferentiation as demonstrated by the loss of neuroendocrine features in small cell lung cancer when SMARCA4 is inhibited.^23^ In melanoma, SMARCA4 mutations have also been identified as a marker of poor prognosis in patients with brain metastases and are associated with depletion of immune signaling gene sets.^24^ Given the essential role of SWI/SNF complexes in maintaining chromatin states and regulating cell fates, it is plausible that a mutation in SMARCA4 may have resulted in epigenetic instability predisposing this patient’s tumor to phenotypic plasticity. The RMS phenotype emerged in the context of entinostat, an HDAC inhibitor, which further alters the epigenetic landscape. This may have created an opportunity for the lineage switch observed to occur under the intense selective pressure of immune checkpoint inhibition.

Transcriptomic profiling provided further evidence of the distinct identities of the two lineages. RMS samples showed upregulation of myogenesis pathways and key muscle differentiation markers such as DES, MYL4 and MYLPF and the melanoma samples retained melanocytic markers like SOX10 (**Figure 2A**). While high-plex imaging showed segregated melanoma and RMS cell populations, the transcriptomic profile of the initial liver lesion (MEL_M1) suggest the presence of an intermediate state. Histologically this lesion was identified as melanoma, but the transcriptome was more similar to that of the later RMS lesions. As the profile was derived from bulk sequencing it could potentially represent a mixture of two cell types, but the uniform melanoma histology suggests that it is a plastic state illustrating that phenotypic switching can occur as a continuum. The GSEA showed significant enrichment of the EMT pathway signature in the RMS samples relative to melanoma, a known feature of tumor plasticity and therapy resistance in melanoma.^25^ No significant fusion, including canonical RMS-associated fusions such as PAX-FOXO1 were identified, suggesting that the emergence of the RMS phenotype was due to epigenetic reprogramming rather than a specific genetic event.

Investigating the tumor extrinsic differences between the two phenotypes revealed notable alterations in the tumor microenvironment. While the metastatic samples of both phenotypes had a relatively small quantity of infiltrating lymphocytes, the RMS lesions exhibited a significantly higher proportion of CD163^+^, often referred to as “M2-like”, immunosuppressive macrophages. Given the initial mixed response seen during the emergence of the RMS phenotype, differences in tumor microenvironments may have contributed to the immune evasion seen with the RMS lineage. Notably, in both lineages, high expression of NGFR was described, which indicates the development of a dedifferentiated neural crest-like phenotype. this was not seen in the only pre-entinostat sample (MEL_LN) available for analysis, it is unclear if this occurred through the same mechanism or converged in parallel by different mechanisms in each phenotype.

This case has several important implications. First, it demonstrates a case of trans-differentiation of tumors that are derived from tissues that diverge early in embryological development. While there are case reports describing a similar phenomenon, this is to our knowledge, the first report with phylogenetic analysis demonstrating the common genetic ancestor of these distinct phenotypes and the first case showing distinct rhabdomyosarcoma and melanoma histologies in spatially segregated metastatic lesions.^7–9^ Second, the case highlights phenotypic plasticity as a potential mechanism of immunotherapy resistance. Finally, the case identifies several of the underlying molecular characteristics of this phenotypic change, driven not by acquired mutations, but rather in changes at the transcriptional level while under the selective pressure of immune checkpoint inhibitor therapy.

Our study has several limitations. While we provide evidence of clonal relatedness and divergence, we are unable to identify the precise molecular event that triggered the transition to the RMS phenotype. Chromatin accessibility profiling, such as ATAC-seq, could have provided additional information regarding if the transcriptional changes observed were related to epigenetic modifications, but was not feasible due to sample constraints. The lack of a pre-treatment RMS sample limited direct comparison for some markers like NGFR, necessitating comparison with historical controls. Finally, differences between the different metastatic sites and timepoints cannot be excluded as a form of sampling bias.

In conclusion, this report details a case of acquired immunotherapy resistance mediated by phenotypic plasticity in which metastatic melanoma underwent transdifferentiation into a distinct RMS phenotype. This process was marked by divergent genomic evolution with notable transcriptomic reprogramming potentially related to epigenetic reprogramming in the context of HDAC inhibitor therapy and baseline mutations in chromatin remodeling. These findings demonstrate the dynamic nature of tumor evolution under the selective pressure of immunotherapy and detail a unique mechanism of resistance.

## Data Availability

All data produced in the present study are available upon reasonable request to the authors

## Acknowledgements

We would like to thank funding support from the Dana Farber Cancer Institute / Harvard CancerCare GI SPORE Career Enhancement Award (AM), Sky Foundation Pancreatic Cancer Research Grant (AM), Doris Duke Charitable Foundation Physician Scientist Fellowship (AM), DF/HCC K12 (K12CA087723) Paul Calabresi Award for Clinical Oncology (AM), NIH K08 (K08CA234458) (DL) and Harvard Ludwig Center (JL and PKS)

## Methods

### Patient Samples

IRB approval was obtained prior to study enrollment and written informed consent was obtained from the patient for the collection of tissue and blood samples for research and genomic profiling, as approved by authors’ Institutional Review Board. Samples were collected and processed at the authors’ institution. Plasma and PBMCs were derived from 10– 30cc whole blood collected in Mononuclear Cell Preparation Tubes. Plasma and PBMCs were isolated according to the BD manufacturer protocol and stored at −80C.

### Immunohistochemistry, H&E, Pathology

Whole slide 5 μm sections were stained with S100 (clone EP32, ready-to-use (RTU), Leica Biosystems, Wetzlar, Germany RTU), SOX10 (clone SP267, RTU, Roche Diagnostics, Basel, Switzerland), desmin (clone DE-R-11, RTU, Leica Biosystems), concentration: RTU), myogenin (clone LO26, RTU, Leica Biosystems), myoD1 (clone EP212, RTU Roche Diagnostics) on Leica Bond III automated stainer (Leica Biosystems).

### DNA/RNA Extraction and Whole Exome Sequencing (WES)

DNA extraction, whole exome library prep and sequencing was performed for samples as previously described.^26,27^ Slides were cut from FFPE blocks and examined by a board-certified pathologist to select high-density cancer foci and ensure high purity of cancer DNA. Biopsy cores were taken from the corresponding tissue block for DNA/RNA extraction. DNA and RNA extraction was performed using Qiagen AllPrep DNA/RNA Mini Kit (#51306), and stored at −20 degrees Celsius. Whole exome capture libraries were constructed from 100ng of DNA from tumor and normal tissue after sample shearing, end repair, and phosphorylation and ligation to barcoded sequencing adaptors. Ligated DNA was size selected for lengths between 200-350 bp and subjected to exonic hybrid capture using Illumina library preps. The sample was multiplexed and sequenced using Illumina HiSeq technology. The Illumina exome uses Illumina’s in-solution DNA probe based hybrid selection method that uses similar principles as the Broad Institute-Agilent Technologies developed in-solution RNA probe based hybrid selection method to generate Illumina exome sequencing libraries.^28,29^

Total RNA was assessed for quality using the Caliper LabChip GX2. The percentage of fragments with a size greater than 200nt (DV200) was calculated using software. An aliquot of 200ng of RNA was used as the input for first strand cDNA synthesis using Illumina’s TruSeq RNA Access Library Prep Kit. Synthesis of the second strand of cDNA was followed by indexed adapter ligation. Subsequent PCR amplification enriched for adapted fragments. The amplified libraries were quantified using an automated PicoGreen assay.

200ng of each cDNA library, not including controls, were combined into 4-plex pools. Capture probes that target the exome were added, and hybridized for recommended time. Following hybridization, streptavidin magnetic beads were used to capture the library-bound probes from the previous step. Two wash steps effectively remove any nonspecifically bound products. These same hybridization, capture and wash steps are repeated to assure high specificity. A second round of amplification enriches the captured libraries. After enrichment the libraries were quantified with qPCR using the KAPA Library Quantification Kit for Illumina Sequencing Platforms and then pooled equimolarly. The entire process is in 96-well format and all pipetting is done by either Agilent Bravo or Hamilton Starlet.

Pooled libraries were normalized to 2nM and denatured using 0.2 N NaOH prior to sequencing. Flowcell cluster amplification and sequencing were performed according to the manufacturer’s protocols using either the HiSeq 2000 v3 or HiSeq 2500. Each run was a 76bp paired-end with a dual eight-base index barcode read. Data was analyzed using the Broad Picard Pipeline which includes de-multiplexing and data aggregation.

### Computational Analysis of WES data

#### Quality control and variant calling

Initial exome sequence data processing and analysis were performed using a customized version of the Getz Lab WES analysis pipeline at the Broad Institute. This pipeline can be found here: https://portal.firecloud.org/#methods/getzlab/CGA_WES_Characterization_Pipeline_v0.1_Dec2018/. After alignment from the Broad Picard Pipeline, BAM files were uploaded into the Terra infrastructure (https://app.terra.bio) which managed intermediate analysis files executed by analysis pipelines.

Out of an initial 9 samples (8 tumor + 1 blood normal), all passed coverage (> 50x mean target coverage) and contamination estimation (< 5%) thresholds.^30^ We removed 1 tumor due to low tumor purity (< 10% tumor cells and no matched mutations in significantly mutated genes), yielding 7 total tumor samples + 1 matched blood normal for analysis.^12,31^

The MuTect algorithm was applied to identify somatic single-nucleotide variants in targeted exons.^32^ Strelka was applied to identify small insertions or deletions.^33^ Alterations were annotated using Oncotator.^34^ Filters were applied to detect and remove known artifacts and germline variants, including DNA oxidation during sequencing.^35^

#### Copy number variants

Total copy number alterations for individual tumors were inferred using adaptations of a binary segmentation algorithm (CapSeg) comparing fractional exon coverage for tumor segments to a panel of normal samples, generating exomic segments and segment copy number.^36,37^ Copy number data were inspected visually and manually for focal amplifications and deletions, and genes were annotated with Oncotator.^34^ For allelic copy numbers, heterozygous SNPs were identified and integrated with the binary segmentation algorithm (Allelic CapSeg), and further adjusted for tumor purity and ploidy. We then called allelic amplifications and deletions, following previously described criteria integrating segment focality and the revised allelic copy number.^38^

#### Purity and ploidy

Purity and ploidy was estimated using the ABSOLUTE algorithm, which integrates variant allele frequency distributions and copy number variants to estimate absolute tumor purity and ploidy and infer cancer cell fraction (CCF), the proportion of cancer cells in the sample which contain each mutation.^39^ Post-purity and ploidy corrected allelic segments were used to estimate allelic copy number estimates.

#### Comutation plot generation

For visualization of the purity and other patient tumor sample characteristics and some of the key copy number variants and driver mutations, we used the comut package.^40^ To narrow down the list of genes to prioritize for visualization, we visualized those genes with pan-cancer “PANCAN” relevance that showed up in at least one of our samples, using the list from a recent comprehensive characterization of cancer driver mutations and genes.^41^ For CNV visualization, we chose those with known functional relevance either in a pan-cancer context or the context of skin cancers and melanoma more specifically.

#### Phylogenetic analysis

Two complementary approaches were taken to perform phylogenetic analyses: PyClone and PhylogicNDT.^42,43^ A comprehensive list of all called point mutations and small insertion-deletions found in any tumor sample was generated. For each tumor sample, the number of alt and reference reads, estimated CCF, and purity and ploidy corrected minor and major copy number at each mutation locus was generated. PyClone (V0.13.1), a Bayesian clustering method for grouping mutations into clonal structures accounting for tumor purity and allelic copy numbers, was then used to generate clusters of mutations and their estimated CCF for each sample given the described mutation inputs and tumor purity.^43^

The following default parameters were used:

~~~
base_measure_params: {alpha: 1, beta: 1}
beta_binomial_precision_params:
 prior: {rate: 0.001, shape: 1.0}
 proposal: {precision: 0.01}
 value: 1000
concentration:
 prior: {rate: 0.001, shape: 1.0}
 value: 1.0
density: pyclone_beta_binomial
init_method: disconnected
num_iters: 10000
~~~

69 clusters were inferred. After filtering for clusters with at least 3 mutations, 19 clusters remained (**Supplementary Fig. 2**), of which 6 were at a CCF ∼0 for all samples and likely represent artifacts (C2, C6, C10, C11, C15, C16).

Upon analysis of the data, three informative patterns emerged:

1. Clusters with CCF ∼1 in all tumors (C0, C1, C3, C18): Four clusters representing a total of 954 mutations were found at >= 0.6 CCF in all samples (**Supplementary Fig. 3**), and likely collectively represent the ancestral clone.
2. A cluster with CCF ∼0 in melanoma phenotype tumors and present at a CCF of 0.5-1 in rhabdomyosarcoma phenotype tumors (C7) with mutations all found in the same chromosomal segment suggest a common ancestor of the rhabdomyosarcoma phenotype tumors where these mutations were lost through the deletion of that segment of the chromosome. They may have some functional relevance to the phenotypic difference between the lineages if those mutations somehow contribute to the rhabdomyosarcoma phenotype, or if the loss event is preventative of the emergence of the rhabdomyosarcoma phenotype for other reasons (**Supplementary Fig. 4**). There is also a cluster with CCF ∼1 in melanoma phenotype tumors and ∼0 in rhabdomyosarcoma phenotype tumors (C7, **Supplementary Fig. 5**), with mutations found in the same chromosomal segment, suggesting a common ancestor of the rhabdomyosarcoma phenotype tumors with deletion of the chromosomal segment containing the cluster mutations for tumors with inferred CCF of 0.
3. A cluster with CCF ∼1 in melanoma phenotype tumors with mutations scattered across different genomic regions that is not present (CCF ∼0) in rhabdomyosarcoma phenotype tumors (C14, **Supplementary Fig. 6**), suggests the split of the rhabdomyosarcoma lineage ancestor before the accumulation of those mutations by the melanoma lineage ancestor. And another cluster with three mutations in different regions (C62, **Supplementary Fig. 7**) with some mutations shared between the first melanoma metastases, suggests a potentially early split between the clone that ended up becoming the late bone metastasis and the clones that were seen earlier (although we can’t be totally confident in this result, based as it is on only a few mutations).

Hierarchical clustering was performed using seaborn’s *clustermap* method^44–46^ (default clustering using Euclidean distance and the Nearest Point Algorithm for linkage between clusters), and two high level lineages were inferred that corresponded to the two tumor phenotypes. This result was consistent whether all the clusters were used, or only those with more than 2 mutations **(Supplementary Fig. 1**). Reassuringly, CNVs (which were not used to generate the lineages) were consistent with the inferred genomic lineages for each phenotype (melanoma or rhabdomyosarcoma), e.g. the lineage corresponding to the melanoma phenotype was inferred by lacking C4 composed of 34 mutations in chromosome 11 (**Supplementary Fig. 4**), and all tumors in this lineage had a corresponding 11 LOH (**Figure 1C**). Similarly, the lineage corresponding to the rhabdomyosarcoma phenotype lacks C7 composed of 43 mutations in a segment of chromosome 19 (**Supplementary Fig. 5**), which is inferred lost in the tumors in the rhabdomyosarcoma lineage (**Supplementary Fig. 1**). Another cluster (C17 – **Supplementary Fig. 8**), seems to be related to a LOH event on chromosome 16 that is subclonal in some rhabdomyosarcoma phenotype samples and appears clonal in RHAB_M2b, although this could be due to sampling a different region of the tumor.

In parallel, we used PhylogicNDT to reconstruct the phylogeny of metastases from DNA sequencing data and inferred cell fractions. PhylogicNDT implements a multidimensional Dirichlet process to jointly estimate the cell population structure and genetic phylogency across all samples, taking copy number profiles, purity values, and joint mutational calls. In our case, we used the posterior distribution on CCF values associated with each mutation (taking into account purity, copy number profiles using ABSOLUTE).^42^ PhylogicNDT Cluster and BuildTree was run on data from selected subsets of samples with the following default parameters: -rb -ni 2000, --seed 0.

Despite differences in the number of inferred mutational clusters (16 total, with 14 with more than 2 mutations), tumors lineages defined by the PyClone/hierarchical clustering approach were reproduced in the phylogenetic tree(s) inferred by PhylogicNDT (**Supplementary Fig. 9**), and melanoma and rhabdomyosarcoma lineage-differentiating mutational clusters related to the same key LOH CNV events were reproduced with PhylogicNDT. Almost all of the 1015 predicted clonally shared mutations are also predicted within the set of 954 from pyclone. Additionally, clusters C9, C7, and C10 in the Phylogic NDT results correspond nearly exactly to C4, C14, and C62 of the pyclone results. Importantly, despite some variation the inferred phylogenetic structure of tumors within each lineage based on the results of this tool would be the same as the pyclone results.

### Computational analysis of RNA-seq data

#### Gene expression and fusion calling

For RNA-seq, we had bam files aligned to the hg38 genome, so in order to make them comparable and harmonizable with the WES analysis which used the hg19 reference, we converted the bams to fastq files with the “bamtofastq” method. We then utilized the “joint_star_alignment_fusion” method documented on Terra (snapshot 10), with a GENCODE v30 annotation lifted over to hg19 coordinates. This is a workflow that does the following: 1) Takes in paired read FASTQs from multiple samples from the same patient (or just one sample) 2) Trims adapter reads from these FASTQs 3) Performs a first pass STAR alignment that is designed to detect splice junctions in all the samples. This results in a junction file that represents called junctions across all the samples. 4) Performs a second pass STAR alignment that uses the called junction file in addition to normal STAR parameters. This improves detection of junctions that appear robustly in some samples but not others and is essentially a forcecalling-like method 5) Performs salmon gene level quantification of each sample 6) Performs fusion calling with STAR-Fusion on each sample, using STAR-Fusion 1.8 in kick start mode.^47,48^ This workflow results in trimmed read fastqs, coordinate sorted bams from the second pass STAR alignment, Salmon quantification for each sample and Fusion calls from STAR-Fusion for each sample.^49^ The STAR-Fusion calls do not show any of the canonical fusions previously associated with rhabdomyosarcoma, and there is a lack of robust fusions shared across samples so no further analysis of the fusion genes was carried out.

#### Differential gene expression

The STAR quality control metrics from the logs were visualized to examine bulk RNA-seq sample quality (Supplementary Table 2). All samples except for one of the replicates from the liver sample and the bone metastasis (MEL_M1_A and MEL_M3) had >85% mapped reads. The aforementioned samples were of poorer quality across multiple metrics. For example, they had only had 61.63% and 52.77% uniquely mapped reads respectively and were therefore excluded from downstream analysis. Some quality control metrics are reported in **Supplementary Table 2**. The samples from the original lymph node primary (MEL_LN_A and MEL_LN_B) were also excluded due to the fact they were coextracted with DNA (as were all samples) and the WES analysis of the tumor purity from the DNA was that there was not enough tumor to estimate the mutations for this sample. Therefore, this sample was thought to not contain enough tumor to analyze the expression differences between this sample and the other tumor samples. Salmon TPM quantifications were then used for differential gene analysis. Because some samples had replicates and some not, we choose to aggregate the replicates for those samples that did have replicates by taking the average of the gene level TPMs across replicates. The data at the replicate level was then log normalized with the np.log2(x + 1) function. T-tests and Wilcoxon rank sum tests were performed to look for statistically significant differences in gene expression between rhabdomyosarcoma phenotype and melanoma phenotype samples using the scipy python packages *scipy*.*stats*.*ttest*_*ind()* function with equal_*var=False* for the t-test and the *scipy*.*stats*.*ranksums()* function for ranked sum statistical tests.^50^ Multiple hypothesis correction was carried out with the *multitest*.*multipletests()* function with *alpha=0.1*. Following multiple hypothesis correction at the gene level with there were no genes with statistically significant differences between melanoma phenotype and rhabdomyosarcoma phenotype tumors, likely due in large part to variance between samples and the lack of replicates for many of the samples. The genes were ranked by average log2 fold change (log2FC) between melanoma and rhabdomyosarcoma phenotype samples, such that the genes with the highest log2FC values were those most enriched in rhabdomyosarcoma phenotype samples, and those with the lowest (negative) log2FC values were those most enriched in melanoma phenotype samples versus rhabdomyosarcoma phenotype samples. The full pre-ranked gene list was then used for gene set enrichment analyses. Boxplots for the top and bottom fifty genes by log2FC were visualized for both melanoma phenotype samples and rhabdomyosarcoma phenotype samples to show the distributions of expression of the genes that were most enriched for each phenotype. A heatmap with hierarchical clustering of the samples was produced with the *sns*.*clustermap*() function of the seaborn package in python by filtering the gene level log2FC by the unadjusted p-values (p-unadjusted<=0.05) to select genes to visualize that were most reliably associated with one phenotype of the other, even if they did not pass multiple hypothesis correction.

#### Gene set enrichment analysis (GSEA) and Enrichr

GSEA was carried out via the GSEA GUI interface and the aforementioned preranked gene list, ranked by average log2FC of genes between rhabdomyosarcoma phenotype samples and melanoma phenotype samples. Preranked GSEA was performed using a curated collection of gene sets consisting of sets from the Hallmark and all the C2 curated gene sets (including the GO pathway collections) in the MSigDB database (v7.5.1 used).^51^ The default settings were used to compute the enrichment scores, with the more conservative scoring approach triggered by setting “*Enrichment statistic* = ‘*classic’*”, based on the reccomended best practices.^51^ Enricher was run with the top 3000 genes ranked by log2FC for each phenotype and the most significantly enriched Cancer Cell Line Encyclopedia, GO Molecular Function and GO Biological process pathways in rhabdomyosarcoma phenotype tumors as compared to melanoma phenotype tumors were visualized with the default method of the tool.^46^

#### Cell type deconvolution from bulk transcriptomic sequencing

We used CIBERSORTx to infer overall expression (OE) of cell type signatures and therefore estimated cell-type proportions within the bulk RNAseq in all RNAseq samples that passed QC as previously described with the Tirosh and LM22 reference matrices (**Supplementary Fig. 10a, 10b, 11**).^45^ The default settings were used as recommended for bulk RNAseq samples. Details of signature derivation and scoring and code based on the Tirosh set is available as previously described.^44^

### Tissue Cyclic Immunofluorescence (t-CyCIF)

Formalin fixed and paraffin embedded (FFPE) slides were cut at 5 µm thickness and mounted on a glass slide at the authors’ institution’s pathology core. We used tissue cyclic immunofluorescence (t-CyCIF) for multiplexed immunofluorescence and the detailed protocol could be found in Lin et. al.^16^ In brief, the unstained slides were dewaxed and antigen-retrieved on Leica BOND RX as described previously. After preprocessing steps on BOND, the slides were bleached with hydrogen peroxide solution (4.5% H2O2, 25mM NaOH in PBS) to reduce endogenous fluorescence/autofluorescence from tissues. Next, slides were incubated with secondary antibodies (Alexa-647 anti-mouse Invitrogen #A-21236; Alexa-555 anti-goat Invitrogen #A-21432; and Alexa-488 anti-rabbit #A-11034) with 1ug/ml Hoechst 33342 (Invitrogen #A-11034) in Odyssey buffer (LI-COR, Cat. 927401) for blocking The antibodies indicated were diluted in Odyssey buffer and incubated in 4 °C overnight. The detailed antibodies used for t-CyCIF in this study can be found in Supplementary Table 3. After washing 5 times with PBS, slides were mounted with 50% Glycerol and coverslipped. The slides were then imaged with CyteFinder imager (RareCyte, Seattle WA) using 20x objectives with 2×2 binning settings. Following imaging, fluorophores were inactivated in 4.5% H2O2 and 25 mM NaOH in PBS for 1 hour at RT in the presence of white LED light and washed 5 times in PBS. The scanned images were then processed with MCMICRO pipeline as described (https://doi.org/10.1038/s41592-021-01308-y) to obtain intensity values for single cells. Antibodies used listed in Supplementary Table 3.

#### CyCIF data analysis

Single-cell data for the selected markers were analyzed by applying Gaussian mixture model (GMM)–based gating on one-dimensional histograms or two-dimensional scatter plots, followed by manual curation. After gating, single-cell marker positivity tables were generated and subjected to k-means clustering (k = 6) to classify cell populations. The resulting six clusters—Rhab, Mel, Stroma, Lymph, Macro, and Other—were subsequently inspected and assigned based on their marker expression profiles.

## Supplementary Figure Legends

**Supplementary Figure 1:**
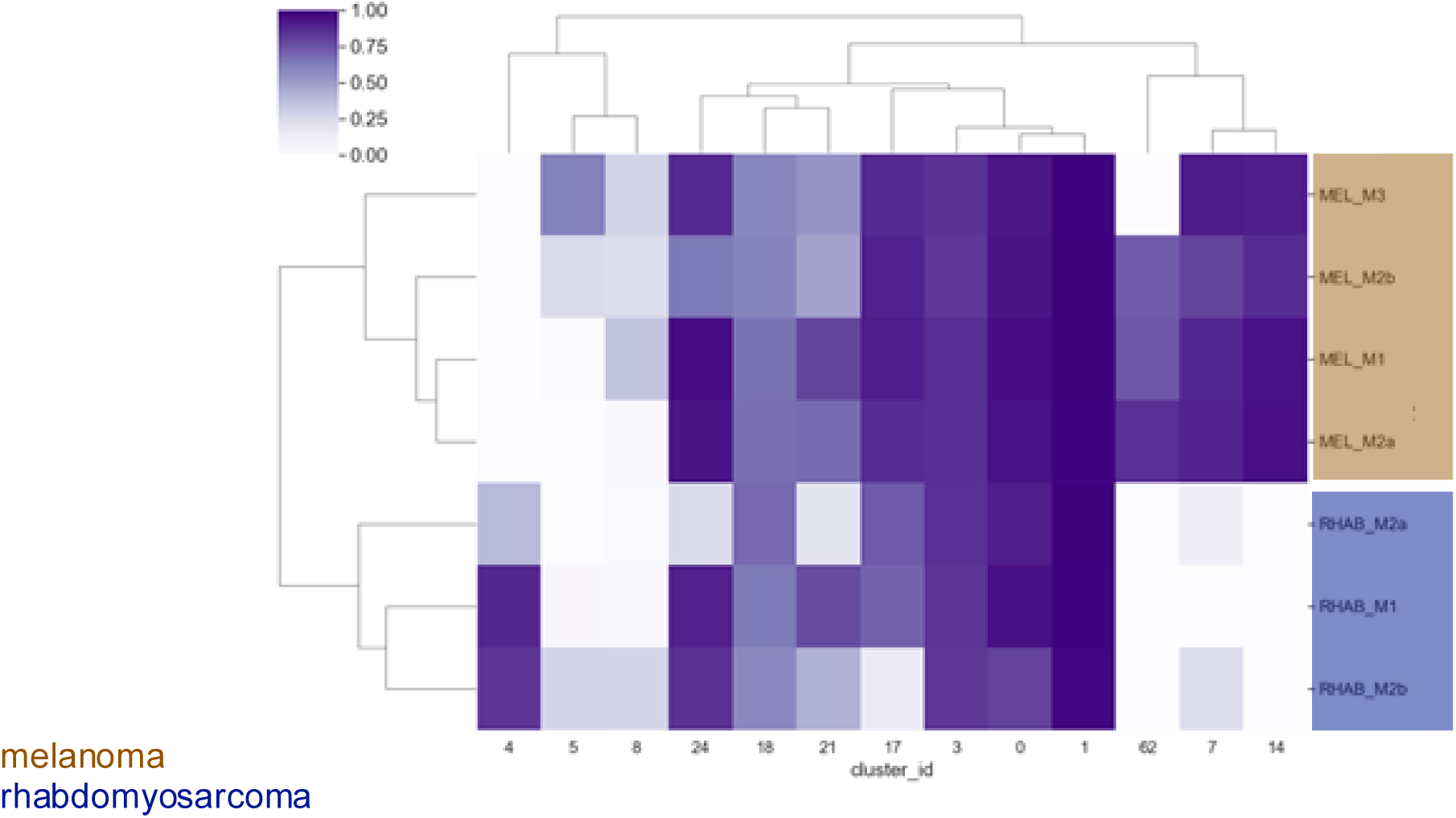
Unsupervised hierarchical clustering of PyClone mutational clusters with at least three mutations also shows clustering by tumor phenotype. Melanoma samples are marked in brown and rhabdomyosarcoma samples marked in dark blue.

**Supplementary Figure 2:**
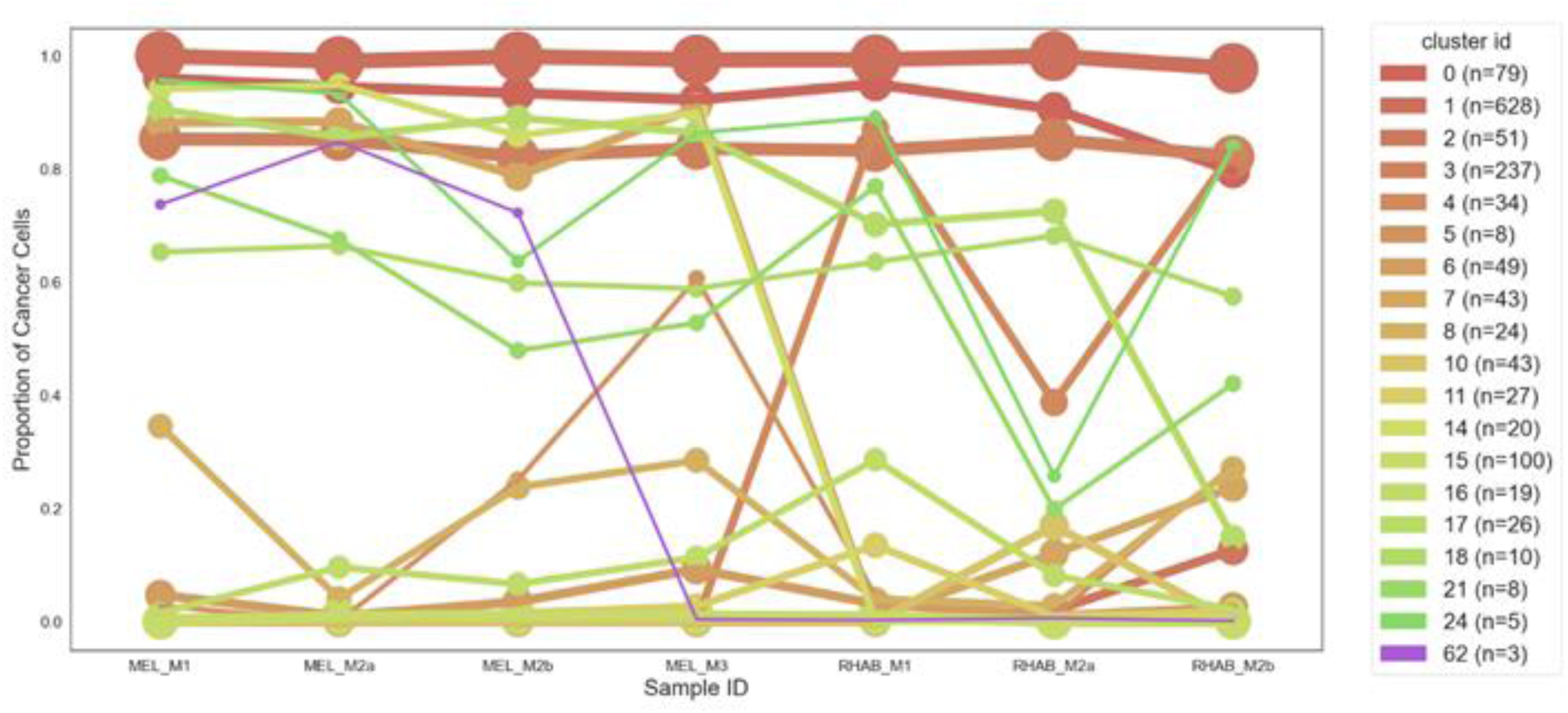
The CCF (cancer cell fraction) in which the mutations are present in each sample is plotted for each sample for all 19 mutational “clusters” with at least three mutations inferred by PyClone

**Supplementary Figure 3:**
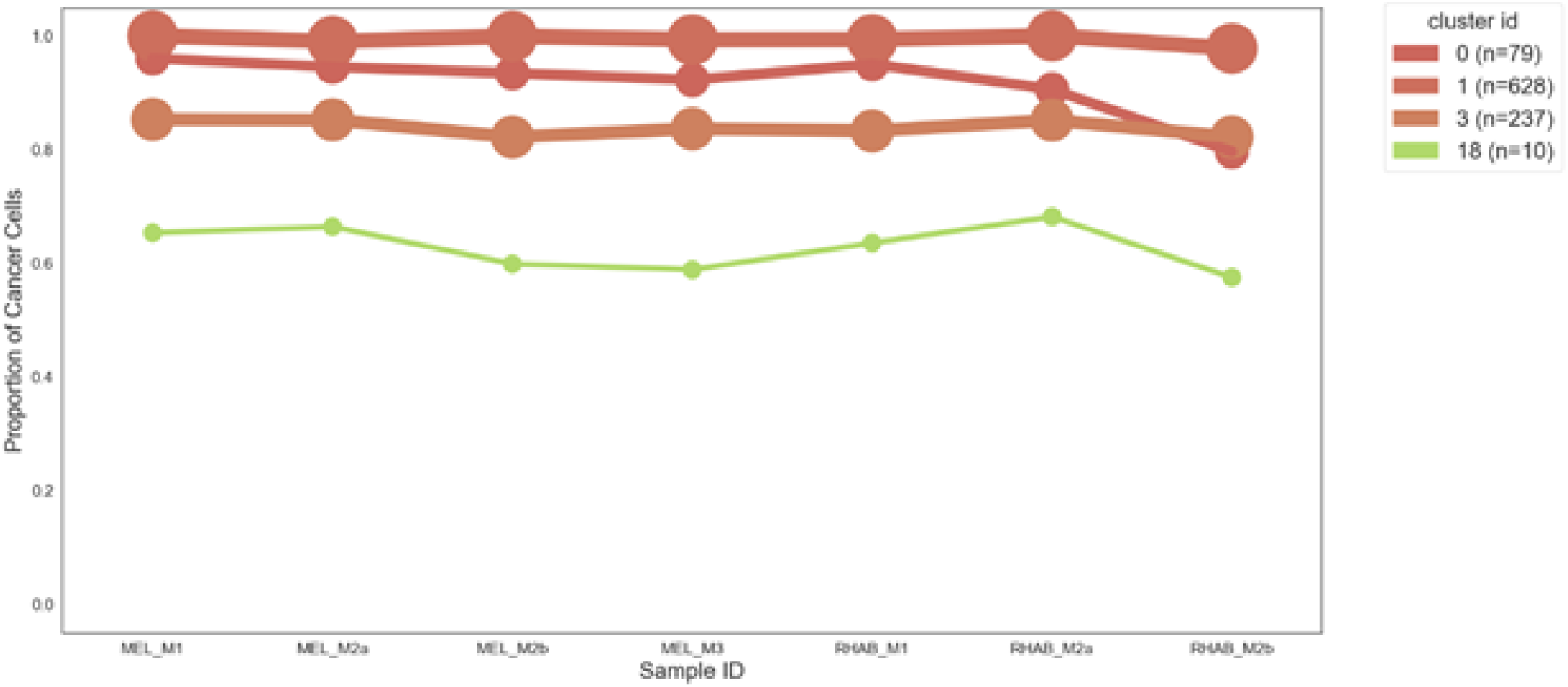
The CCF for all 4 clonally present mutational “clusters” (with at least mutations) inferred by PyClone

**Supplementary Figure 4:**
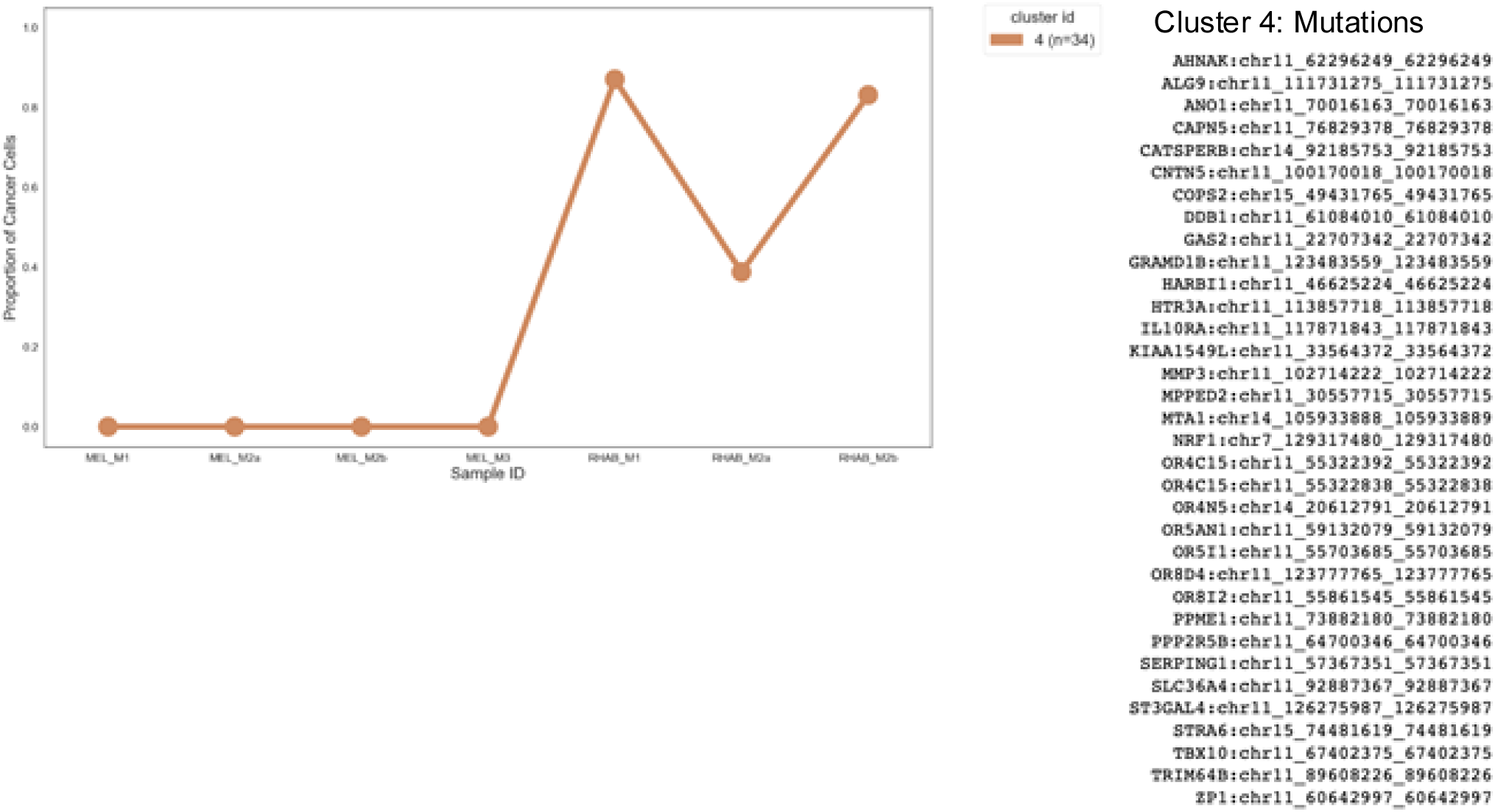
The CCFs (above) across all samples for a mutational cluster (C4 – mutations listed to the right of the graph) in PyClone that seems to represent a loss event (LOH) in chr11 in melanoma samples.

**Supplementary Figure 5:**
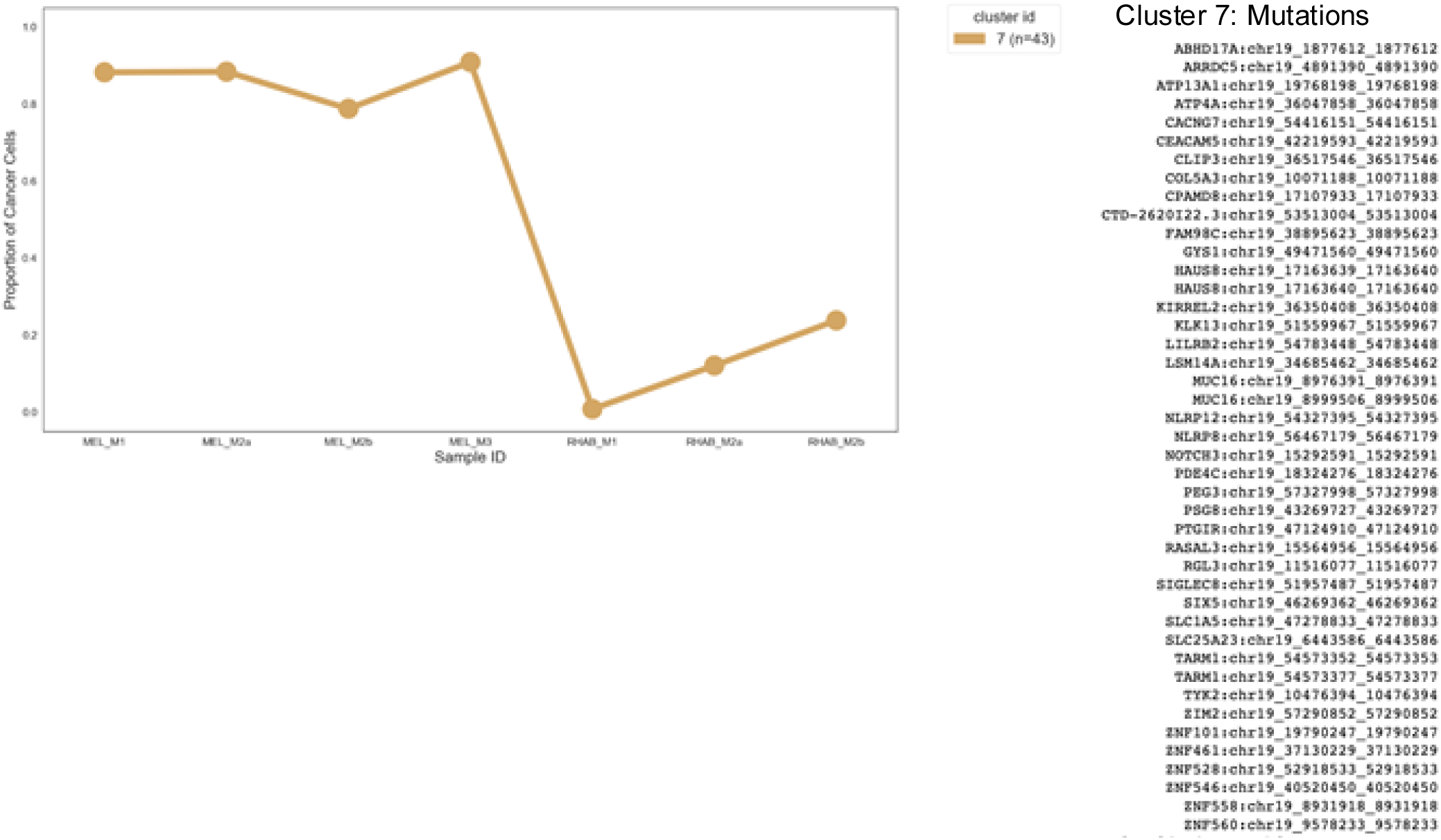
The CCFs (left) across all samples for a mutational clusters (C7 – mutations listed to the right of the graph) in PyClone that seems to represent a loss event (LOH) in chr19 in rhabdomyosarcoma samples.

**Supplementary Figure 6:**
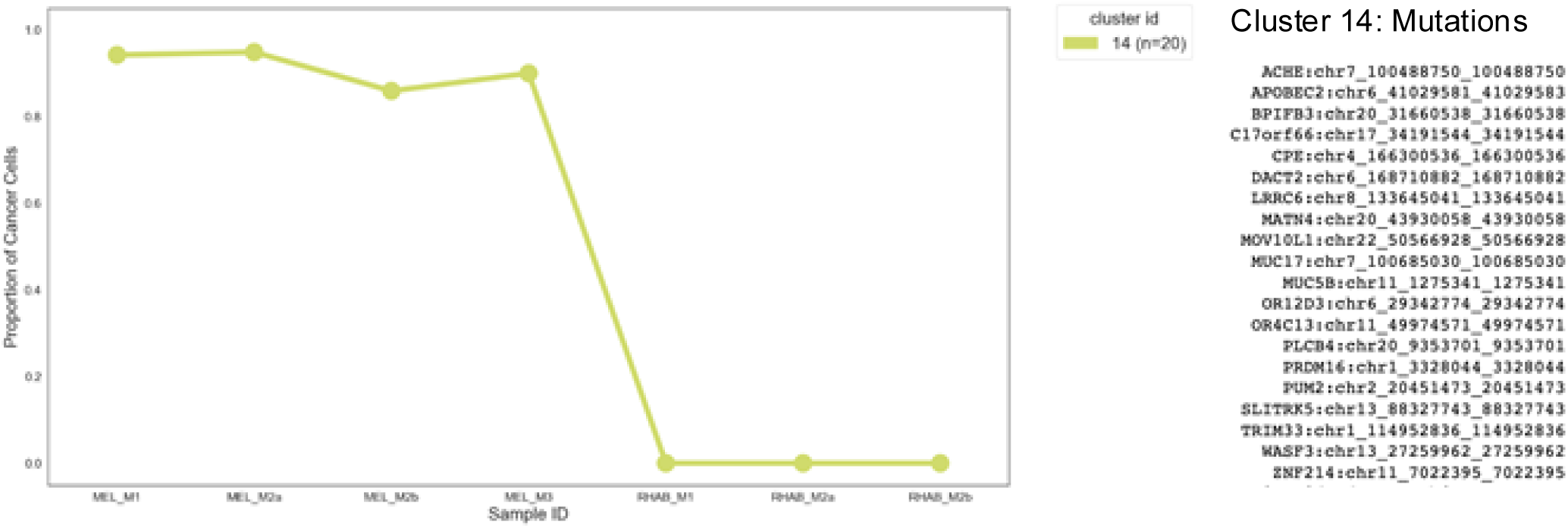
The CCFs (left) across all samples for a mutational clusters (C14 – mutations listed to the right of the graph) in PyClone that seems to represent mutations gained in a common ancestor of the melanoma phenotype samples but after the split with the rhabdomyosarcoma phenotype samples (as they are not seen in those samples.

**Supplementary Figure 7:**
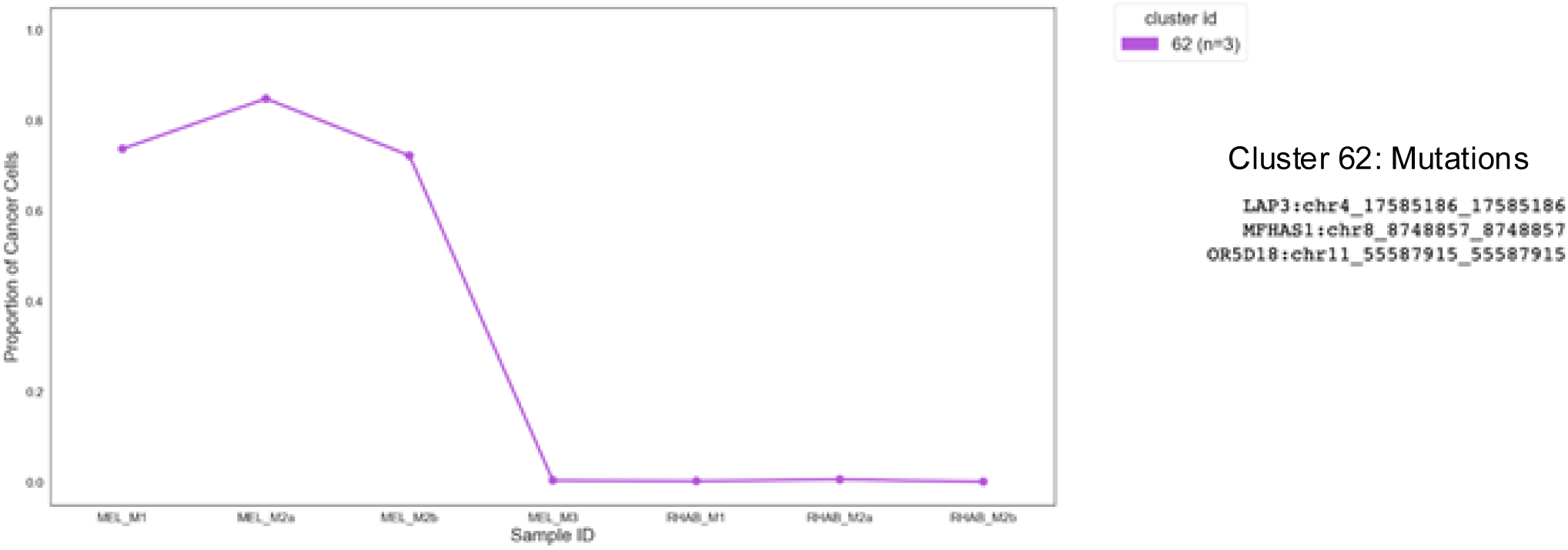
The CCFs (left) across all samples for a mutational clusters (C62) in PyClone that seems to represent mutations gained in an ancestral clone of MEL_M1 and MEL_M2 after the split of the MEL_M3 clone and the rhabdomyosarcoma phenotype samples (as these mutations are not seen in those samples).

**Supplementary Figure 8:**
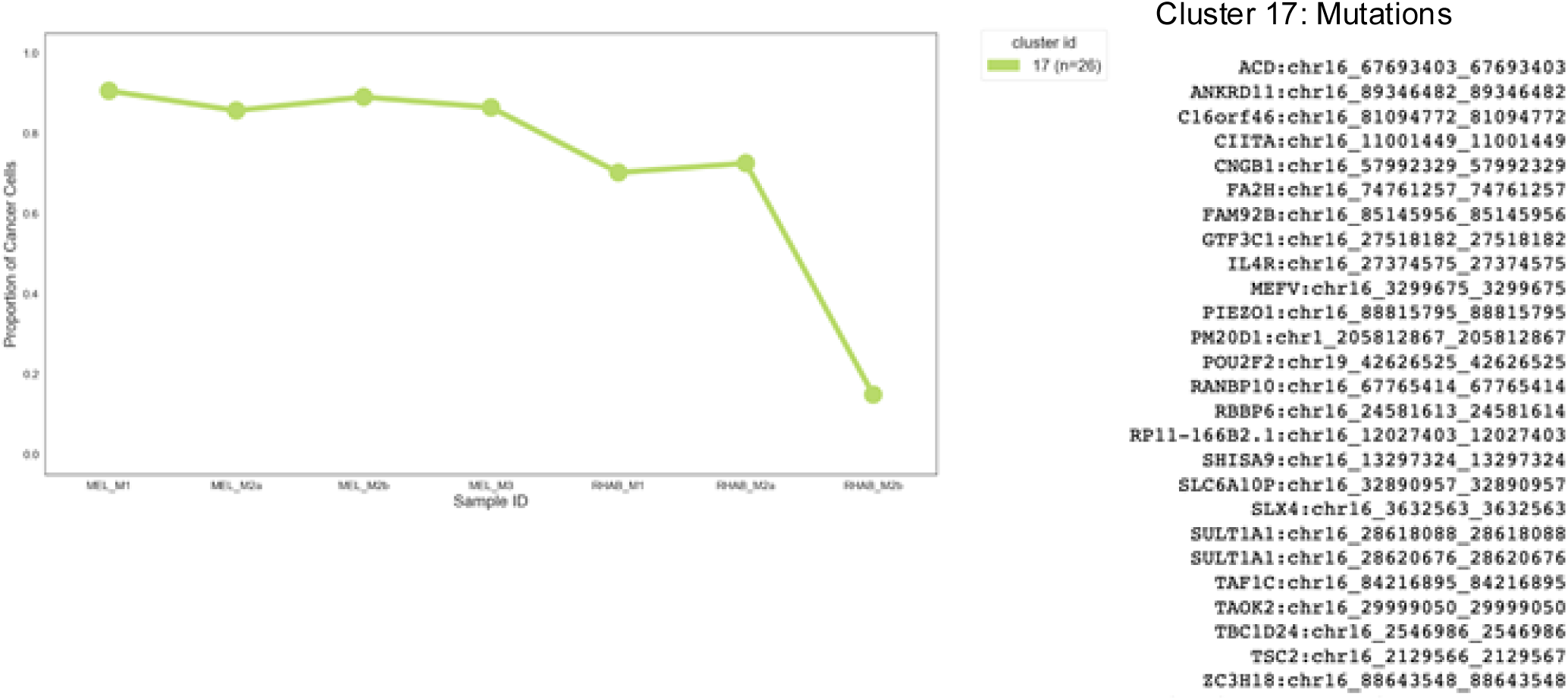
The CCFs (left) across all samples for a mutational clusters (C17 – mutations listed to the right of the graph) in PyClone that seems to represent a loss event (LOH) in chr16 in rhabdomyosarcoma samples that is subclonal (or less sampled) in earlier samples (M1, M2a) and clonal for the M2b sample taken from a different region of the M2 metastasis.

**Supplementary Figure 9:**
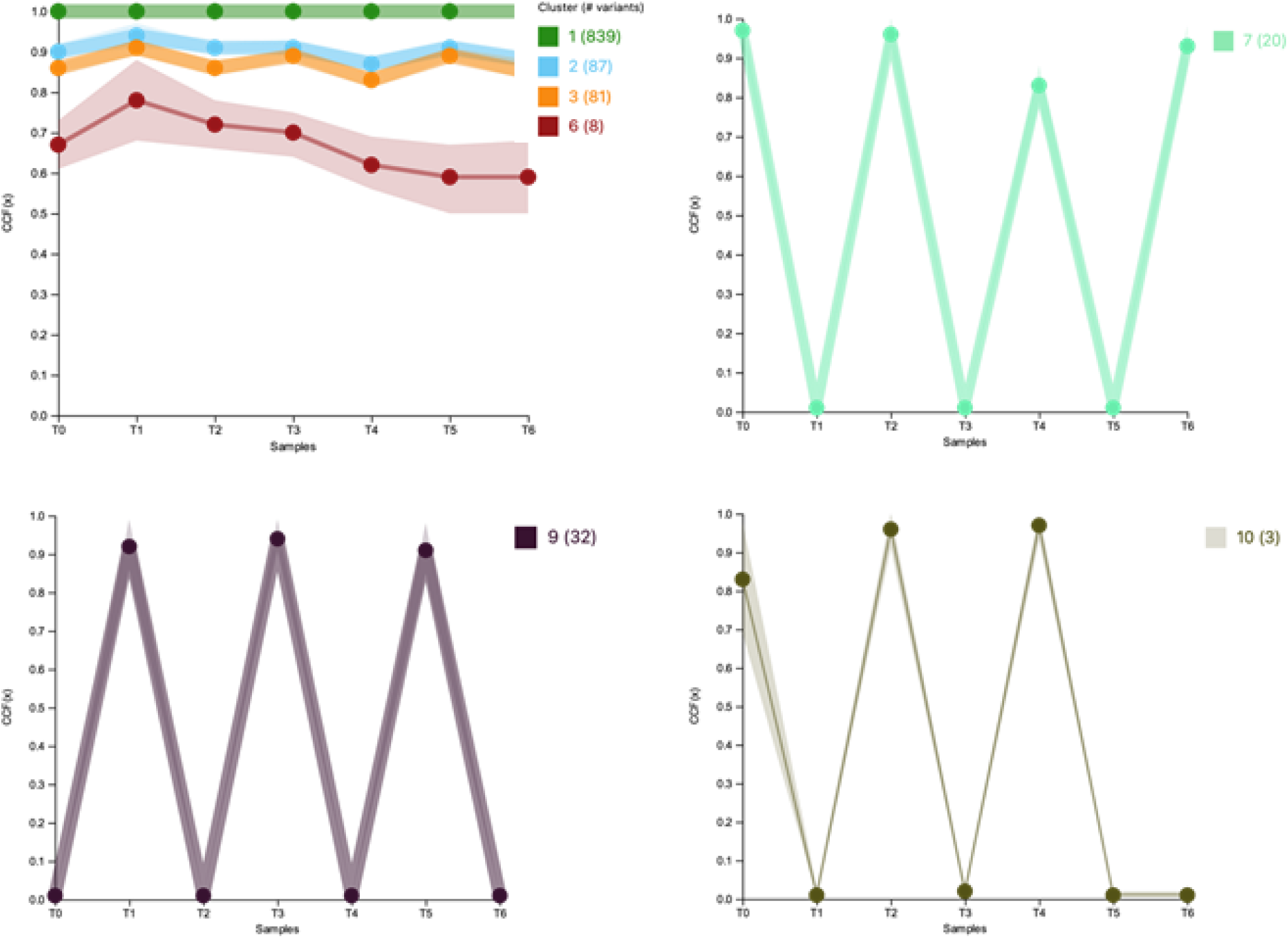
Examples showing CCFs for mutational clusters computed with PhylogicNDT supporting similar phylogenetic results to those computed with PyClone. Clockwise from top left are shown the clusters that correspond to mutations that are clonally shared across; a cluster that corresponds to PyClone cluster 14; a cluster that corresponds to PyClone cluster 62, a cluster that corresponds to PyClone cluster 4. (Note that here the samples are in a different order than the PyClone results. Correspondences are as follows: T0 = MEL_M1, T1 = RHAB_M1, T2 = ME_M2a, T3 = RHAB_M2a, T4 = MEL_M2b, T5 = RHAB_M2b, T6 = MEL_M3).

**Supplementary Figure 10a:**
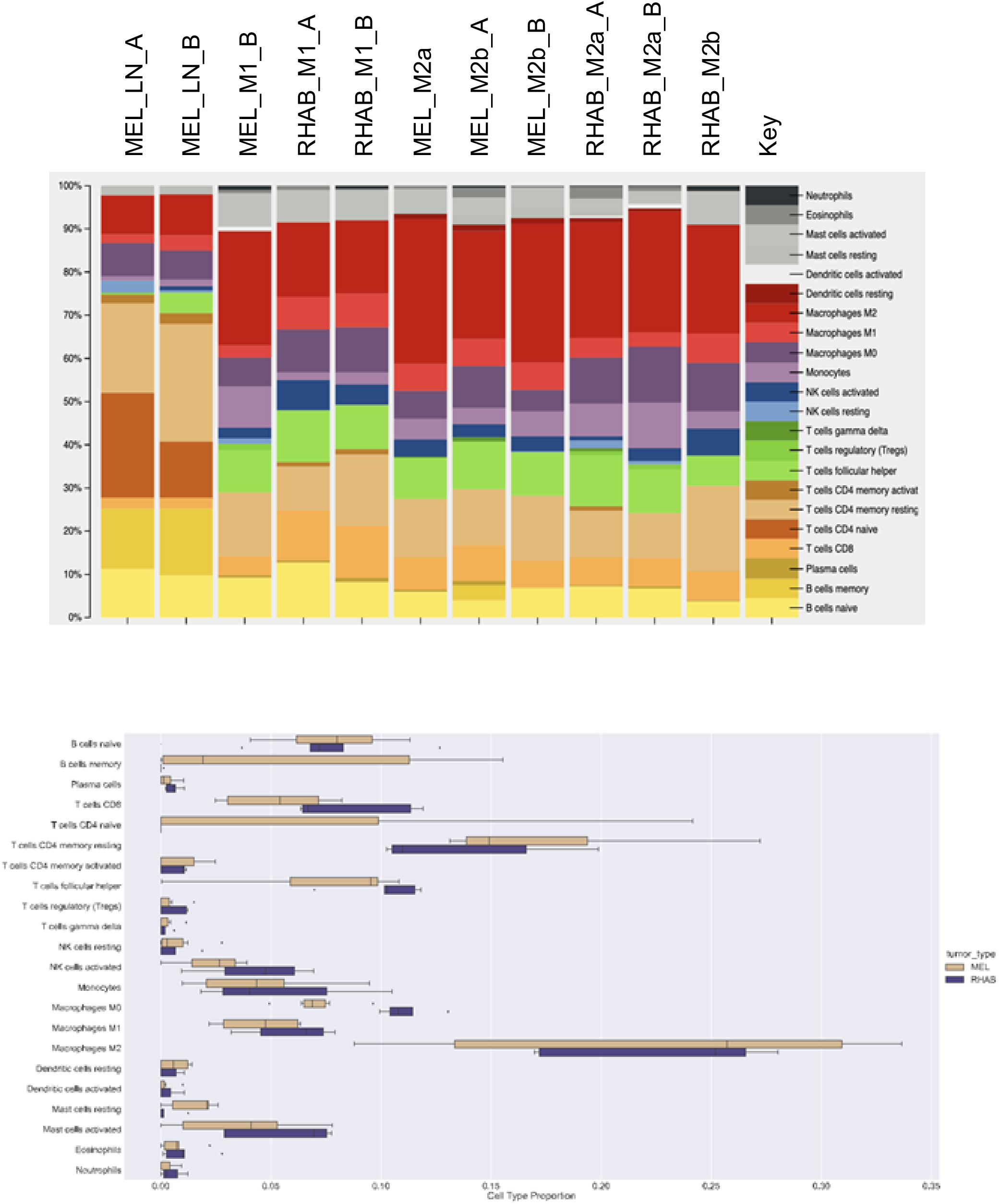
Cell type proportion estimates from CIBERSORTx with the LM22 reference matrix. The distributions of cell type proportion estimates are visualized in the box plot (bottom), and the individual tumor estimates are shown in the stacked bar graph (top)

**Supplementary Figure 10b:**
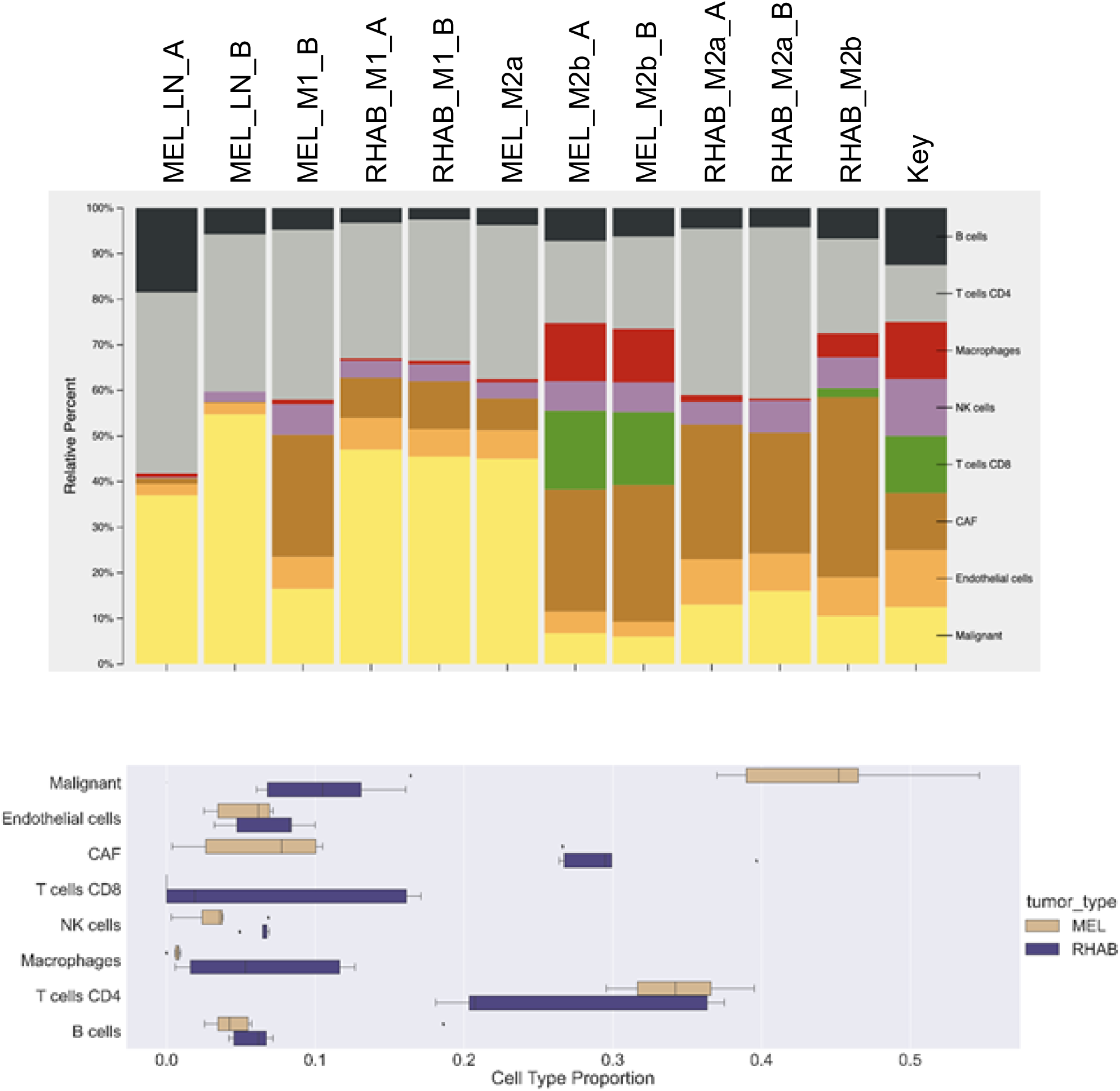
Cell type proportion estimates from CIBERSORTx with the Tirosh reference matrix. The distributions of cell type proportion estimates are visualized in the box plots (bottom), and the individual tumor estimates are shown in the stacked bar graph (top)

**Supplementary Figure 11:**
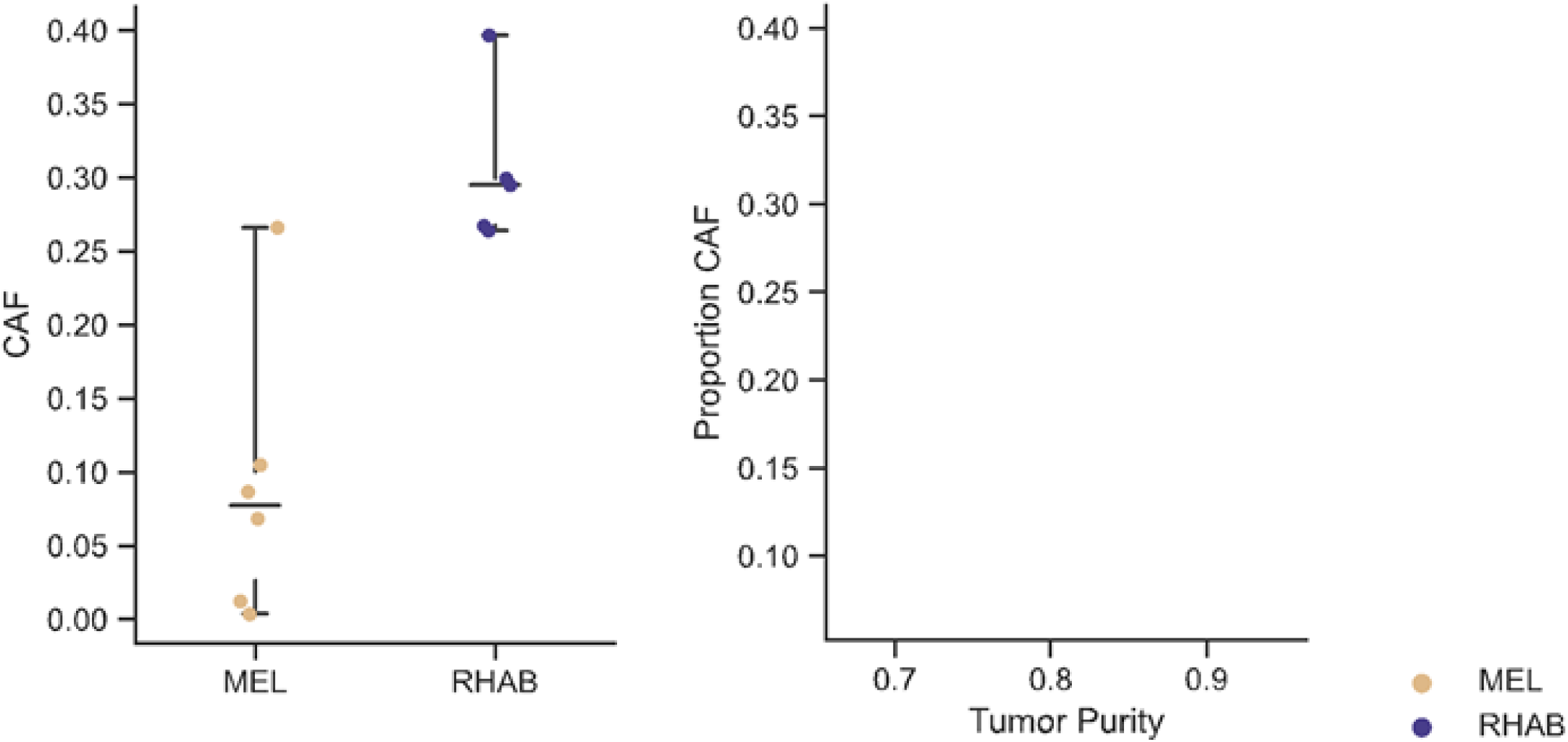
CAF proportions as estimated by CIBERSORTx with the Tirosh reference by tumor type (left), CAF estimates and tumor purity and not obviously associated with each other for these samples

## Supplementary Table Legends

**Supplementary Table 1:**
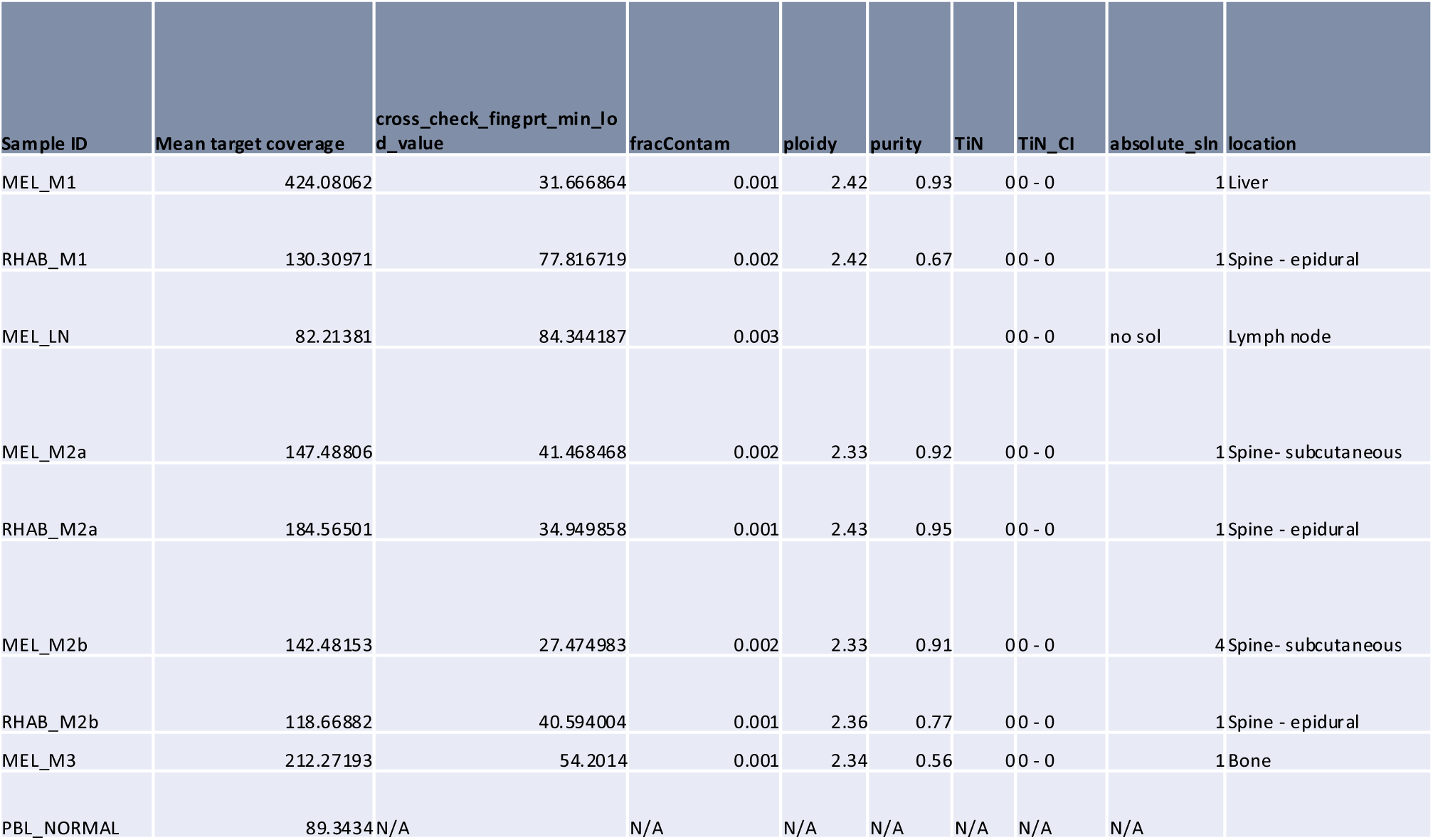
Whole exome sequencing sample quality information.

**Supplementary Table 2:** STAR quality control metrics.

| Sample ID | Uniquely mapped reads % | Mismatch rate per base, % | % of reads mapped to multiple loci | % of reads mapped to too many loci | % of reads unmapped: too many mismatches | % of reads unmapped: too short | % of reads unmapped: other | % of chimeric reads |
| --- | --- | --- | --- | --- | --- | --- | --- | --- |
| MEL_M2a | 87.33 | 0.21 | 10.73 | 0.09 | 0.06 | 1.64 | 0.15 | 1.12 |
| MEL_LN | 87.95 | 0.21 | 9.3 | 0.16 | 0.04 | 2.28 | 0.27 | 1.15 |
| MEL_M1 | 87.8 | 0.21 | 9.77 | 0.26 | 0.06 | 1.88 | 0.24 | 1.2 |
| RHAB_M1 | 89.35 | 0.2 | 8.41 | 0.19 | 0.04 | 1.7 | 0.31 | 1.05 |
| RHAB_M2b | 90.01 | 0.3 | 7.26 | 0.42 | 0.17 | 1.55 | 0.57 | 1.01 |
| RHAB_M2a | 86.79 | 0.2 | 9.91 | 0.16 | 0.04 | 2.87 | 0.22 | 1.91 |
| MEL_M3 | 52.77 | 0.72 | 29.71 | 11.47 | 2.05 | 2.65 | 1.34 | 3.87 |
| MEL_M2a | 88.02 | 0.21 | 9.21 | 0.1 | 0.05 | 2.47 | 0.15 | 1.81 |
| MEL_M2b | 89.58 | 0.29 | 8.03 | 0.18 | 0.17 | 1.81 | 0.23 | 1.36 |
| RHAB_M2a | 87.47 | 0.24 | 9.81 | 0.14 | 0.05 | 2.34 | 0.18 | 1.9 |
| MEL_LN | 89.19 | 0.22 | 8.91 | 0.13 | 0.06 | 1.5 | 0.21 | 0.69 |
| MEL_M1 | 61.63 | 0.38 | 32.78 | 1.54 | 0.48 | 3.27 | 0.31 | 3.34 |
| RHAB_M1 | 87.29 | 0.22 | 10.65 | 0.18 | 0.04 | 1.56 | 0.29 | 0.82 |

**Supplementary Table 3:** Antibodies used for CyCIF.

| Cycle | Channel | Target | Vendor | Catalog | Host | Conjugated | Clone | Function | Dilution |
| --- | --- | --- | --- | --- | --- | --- | --- | --- | --- |
| Cycle1 | FITC/488 | anti-CD3 | Abcam | ab11089 | Rat | n/a | CD3-12 | Immune | 1:400 |
|  | Cy3/555 | anti-S100 | DAKO | Z0311 | Rabbit | n/a | Poly | Tumor | 1:400 |
|  | Cy5/647 | anti-CD8 | DAKO | M7103 | Mouse | n/a | C8/144B | Immune | 1:100 |
| Cycle2 | FITC/488 | CD45RO | Biolegend | 304212 | Mouse | Alexa 488 | UCHL1 | Immune | 1:200 |
|  | Cy3/555 | panCK | Ebio | 41-9003-82 | Mouse | eFluor 570 | AE1/AE3 | Tumor | 1:600 |
|  | Cy5/647 | SMA | Ebio | 50-9760-82 | Mouse | Alexa 647 | 1A4 | Stroma | 1:800 |
| Cycle3 | FITC/488 | Ki-67 | CST | 11882S | Rabbit | Alexa 488 | D3B5 | Other | 1:400 |
|  | Cy/555 | CD68 | CST | 79594S | Rabbit | Alexa 555 | D4B9C | Immune | 1:400 |
|  | Cy5/647 | PD-L1 | CST | 15005S | Rabbit | Alexa 647 | E1L3N | Immune | 1:200 |
| Cycle4 | FITC/488 | CD4 | Abcam | ab196372 | Rabbit | Alexa 488 | EPR6855 | Immune | 1:200 |
|  | Cy3/555 | CD45 | Biolegend | 304039 | Mouse | PE | HI30 | Immune | 1:100 |
|  | Cy5/647 | PD-1 | Abcam | ab201825 | Rabbit | Alexa 647 | EPR4877(2) | Immune | 1:200 |
| Cycle5 | FITC/488 | CD163 | Abcam | ab218293 | Rabbit | Alexa 488 | EPR14643-36 | Immune | 1:200 |
|  | Cy3/555 | FOXP3 | Ebio | 41-4777-80 | Mouse | eFluor 570 | 236A/E7 | Immune | 1:100 |
|  | Cy5/647 | CD31 | Abcam | ab218582 | Rabbit | Alexa 647 | EPR3094 | Stroma | 1:600 |
| Cycle6 | FITC/488 | EGFR | CST | 5616S | Rabbit | Alexa 488 | D38B1 | Other | 1:200 |
|  | Cy3/555 | Vimentin | CST | 9855S | Rabbit | Alexa 555 | D2H3 | Tumor | 1:1200 |
|  | Cy5/647 | NGFR | BD | 580326 | Mouse | Alexa 647 | C40-1457 | Other | 1:200 |
| Cycle7 | FITC/488 | MITF | abcam | ab201675 | Rabbit | Alexa 488 | D5 | Tumor | 1:400 |
|  | Cy3/555 | H3K27Me3 | CST | 40724S | Rabbit | PE | C36B11 | Other | 1:200 |
|  | Cy5/647 | S100A4 | Abcam | ab196168 | Rabbit | Alexa 647 | EPR2761(2) | Stroma | 1:400 |
| Cycle8 | FITC/488 | E-Cadherin | CST | 3199S | Rabbit | Alexs 488 | 24E10 | Tumor | 1:400 |
|  | Cy3/555 | Desmin | Abcam | ab203422 | Rabbit | Alexa 555 | Y66 | Stroma/<br>Tumor | 1:1000 |
|  | Cy5/647 | SOX10 | Abcam | ab270151 | Rabbit | Alexa 647 | SP267 | Tumor | 1:300 |

